# Cumulative genetic risk and *C9orf72* repeat status independently associate with ALS status in two case-control studies

**DOI:** 10.1101/2022.10.27.22281377

**Authors:** John F. Dou, Kelly M. Bakulski, Kai Guo, Junguk Hur, Lili Zhou, Sara Saez-Atienzar, Ali R Stark, Ruth Chia, Alberto García-Redondo, Ricardo Rojas-García, Juan Francisco Vázquez-Costa, Ruben Fernandez Santiago, Sara Bandres-Ciga, Pilar Gómez-Garre, Maria Teresa Periñán, Pablo Mir, Jordi Pérez-Tur, Fernando Cardona, Manuel Menendez-Gonzalez, Javier Riancho, Daniel Borrego-Hernández, Lucía Galán-Dávila, Jon Infante Ceberio, Pau Pastor, Spanish Neurological Consortium, Bryan J. Traynor, Eva L. Feldman, Stephen A Goutman, Members of the Spanish Neurological Consortium:, Jesús Esteban-Pérez, Pilar Cordero-Vázquez, Sevilla Teresa, Adolfo López de Munain, Julio Pardo-Fernández, Ivonne Jericó-Pascual, Oriol Dols-Icardo, Ellen Gelpi Mantius, Janet Hoenicka, Victoria Alvarez Martinez, Francisco Javier Rodríguez de Rivera Garrido, Katrin Beyer, Jordi Clarimón Echevarría

**Affiliations:** Department of Epidemiology, School of Public Health, University of Michigan, Ann Arbor, MI; Department of Neurology, University of Michigan, Ann Arbor, MI; NeuroNetwork for Emerging Therapies, University of Michigan, Ann Arbor, MI; Department of Biomedical Sciences, University of North Dakota, Grand Forks, ND; Department of Biostatistics, School of Public Health, University of Michigan, Ann Arbor, MI; Neuromuscular Diseases Research Section, Laboratory of Neurogenetics, National Institute on Aging, National Institutes of Health, Bethesda, MD 20892, USA; ALS Unit, Instituto de Investigación Sanitaria ‘i + 12’ del Hospital Universitario 12 de Octubre de Madrid, SERMAS, Madrid, Spain; CIBERER, Center for Networked Biomedical Research into Rare Diseases, Madrid, Spain; Neuromuscular Disorders Unit, Neurology Department and Sant Pau Biomedical Research Institute, Hospital de la Santa Creu I Sant Pau, Universitat Autonoma de Barcelona, Barcelona, Spain; Neuromuscular Unit, Hospital Universitario y Politécnico la Fe, IIS La Fe, Valencia, Spain; Department of Medicine, Universitat de València, Valencia, Spain; Centro de Investigación Biomédica en Red sobre Enfermedades Neurodegenerativas (CIBERNED), Madrid, Spain; Lab of Parkinson’s disease and Other Neurodegenerative Movement Disorders, IDIBAPS-Institut d’Investigacions Biomèdiques, Barcelona, Catalonia, Spain; Unitat de Parkinson i Trastorns del Moviment. Servicio de Neurologia, Hospital Clínic de Barcelona and Institut de Neurociencies de la Universitat de Barcelona (Maria de Maetzu Center), Catalonia, Spain; Center for Alzheimer’s and Related Dementias, National Institute on Aging, Bethesda, Maryland, USA; Unidad de Trastornos del Movimiento, Servicio de Neurología y Neurofisiología Clínica, Instituto de Biomedicina de Sevilla, Hospital Universitario Virgen del Rocío/CSIC/Universidad de Sevilla, Sevilla, Spain; Departamento de Medicina, Universidad de Sevilla, Sevilla, Spain; Neurology and Molecular Genetics Mixed Investigation Unit. Instituto de Investigación Sanitaria La Fe, Valencia, Spain; Molecular Genetics Unit. Institut de Biomedicina de València-CSIC, Valencia, Spain; Department of Medicine, Universidad de Oviedo, Oviedo, Spain; Department of Neurology, Hospital Universitario Central de Asturias, Oviedo, Spain; Instituto de Investigación Sanitaria del Principado de Asturias, Oviedo, Spain; Service of Neurology. Hospital Sierrallana. IDIVAL University of Cantabria, Barrio Ganzo s/n. 39300. Torrelavega. Spain; Instituto de Investigación Marqués de Valdecilla, Santander, Spain; Department of Neurology, ALS Unit, Hospital Clínico Universitario ‘San Carlos’, Madrid, Spain; Unit of Neurodegenerative diseases, Department of Neurology, University Hospital Germans Trias I Pujol, Badalona, Barcelona, Spain; Neurosciences, The Germans Trias i Pujol Research Institute (IGTP) Badalona, Barcelona, Spain; Universitat de Valencia, Valencia, Spain; Neuroscience Area, Institute Biodonostia, and Department of Neurosciences, University of Basque Country EHU-UPV, San Sebastian, Spain; Neurology Department, Hospital Universitario Donostia, San Sebastian, Spain; Neurology Department, Hospital Clinico, Santiago de Compostela, Spain; Neurology Department. Complejo Hospitalario de Navarra, Pamplona, Spain; Department of Neurology. ALS Clinic. Hospital Universitario de Navarra. IdisNa (Instituto de Investigación Sanitaria de Navarra), Navarra, Spain; Memory Unit, Neurology Department and Sant Pau Biomedical Research Institute, Hospital de la Santa Creu I Sant Pau, Universitat Autonoma de Barcelona, Barcelona, Spain; Neurological Tissue Bank of the Biobank-Hospital Clinic-IDIBAPS, Barcelona, Spain; Institute of Neurology, Medical University of Vienna, Vienna, Austria; Laboratory of Neurogenetics and Molecular Medicine-Pediatric Institute of Rare Diseases, Institut de Recerca Sant Joan de Déu, Barcelona, Spain; Laboratorio de Genética, Hospital Universitario Central de Asturias, Asturias, Spain; Instituto de Investigación Sanitaria del Principado de Asturias (ISPA), Asturias, Spain; Department of Neurology, ALS Unit, Hospital Universitario La Paz, Madrid, Spain; Department of Pathology, University Hospital Germans Trias I Pujol, Badalona, Barcelona, Spain

**Keywords:** amyotrophic lateral sclerosis, polygenic risk, polygenic scores, classification

## Abstract

**Background:** Most amyotrophic lateral sclerosis (ALS) patients lack a monogenic mutation. This study evaluates ALS cumulative genetic risk in an independent Michigan and Spanish replication cohort using polygenic scores.

**Methods:** ALS (n=219) and healthy control (n=223) participant samples from University of Michigan were genotyped and assayed for the *C9orf72* hexanucleotide expansion. Polygenic scores excluding the C9 region were generated using an independent ALS genome-wide association study (20,806 cases, 59,804 controls). Adjusted logistic regression and receiver operating characteristic curves evaluated the association and classification between polygenic scores and ALS status, respectively. Population attributable fractions and pathway analyses were conducted. An independent Spanish study sample (548 cases, 2,756 controls) was used for replication.

**Results:** Polygenic scores constructed from 275 single nucleotide polymorphisms had the best model fit in the Michigan cohort. A standard deviation increase in ALS polygenic score associated with 1.28 (95%CI 1.04-1.57) times higher odds of ALS with area under the curve of 0.663 versus a model without the ALS polygenic score (p-value=1×10^−6^). The population attributable fraction of the highest 20^th^ percentile of ALS polygenic scores, relative to the lowest 80^th^ percentile, was 4.1% of ALS cases. Genes annotated to this polygenic score enriched for important ALS pathomechanisms. Meta-analysis with the Spanish study, using a harmonized 132 single nucleotide polymorphism polygenic score, yielded similar logistic regression findings (odds ratio: 1.13, 95%CI 1.04-1.23).

**Conclusion:** ALS polygenic scores can account for cumulative genetic risk in populations and reflect disease-relevant pathways. If further validated, this polygenic score will inform future ALS risk models.

**What is already known on this topic:** Amyotrophic lateral sclerosis (ALS) is a complex neurodegenerative disease caused in part by genetic factors, and methods to account for ALS polygenic disease risk are needed.

**What this study adds:** An ALS polygenic score reflects disease risk in the population and helps ascribe the magnitude of the risk.

**How this study might affect research, practice or policy:** ALS polygenic scores can assign the overall distribution of genetic risk in a population and can be used to screen individuals at higher risk.

## Introduction

Amyotrophic lateral sclerosis (ALS) is a fatal neurodegenerative disease characterized by rapidly progressive muscle weakness and death within 2 to 4 years from symptom onset^1 2^ with 50% of patients manifesting cognitive or behavioral dysfunction.^1 2^ ALS is traditionally divided into familial and sporadic forms. Familial ALS, comprising 10-15% of persons with ALS, includes individuals with a family history of ALS or related diseases, whereas sporadic ALS represents individuals without a family history. Under a monogenic model, a single risk gene is associated with a greater likelihood of developing ALS^3^ or contributes to a distinct phenotypic outcome, such as earlier age of disease onset.^3 4^ Since 1994, over 40 genes have been associated with ALS.^5^ The non-coding chromosome 9 open reading frame 72 (*C9orf72*) hexanucleotide expansion is the most common genetic form of ALS and is observed in 40% of familial and 10% of sporadic ALS in mixed European populations.^6 7^ Superoxide dismutase 1 (*SOD1*), TAR DNA binding protein 43 (*TARDBP*), and fused in sarcoma (*FUS*) are the next most common genes with polymorphism frequencies of around 1% or less in sporadic cases.^8^ Importantly, most ALS patients do not carry a single causative ALS risk gene mutation. Further, heritability estimates of ALS range from around 20% for genome-wide complex trait analysis and 60% for twin data^9^ and ALS shares some genetic risk with other diseases and conditions.^10-12^ It is increasingly clear that many common polymorphisms may contribute a small amount of disease risk.^13^ Since most ALS patients do not have a monogenic cause, it is crucial to understand the genetic contribution to ALS beyond single highly penetrant mutations.

We hypothesize that cumulative genetic risk for ALS can be summarized using polygenic scores. To our knowledge, the utility of a polygenic score for ALS, independent of *C9orf72* status, has not been tested to understand the associations with ALS and ALS risk prediction. The goals of the current study were to develop a genome-wide ALS polygenic score using an independent ALS cohort of participants not previously included in any genome-wide association study (GWAS) and test the score contribution to ALS risk models independently of *C9orf72* status.

## Methods

University of Michigan samples: Study procedures of this Institutional Review Board approved longitudinal case/control study are published (see Supplemental Methods).^14-16^ All participants provided informed consent. DNA extraction, genome-wide genotyping, *C9orf72* repeat expansion assay, and data processing followed published protocols and are presented in Supplementary Methods and **Supplementary Figures 1-3**.

Spanish Neurological Consortium: Participants were recruited across several sites in Spain as previously published^17^ or as part of the ALS Genetic Spanish Consortium (ALSGESCO) as previously published^18^ (see Supplemental Methods). All participants provided informed consent and the study received local ethics board approval. The coordination and use of samples for this publication were approved by the institutional review board of the National Institute on Aging. DNA extraction, genome-wide genotyping, *C9orf72* repeat expansion assay, and processing followed published protocols are presented in Supplementary Methods.

Polygenic scores were developed using published methods^19-21^ and were derived from an independent GWAS of 20,806 ALS cases and 59,804 controls^22^ (See Supplementary Methods). Developed polygenic scores both included and excluded the *C9orf72* region (**Figure 1**). Consistent with Polygenic Score Reporting Standards,^23^ for each SNP included in the polygenic score, the identifier, chromosome, position, weight, and p-value of association with ALS were provided (**Supplementary Table S1**).

**Figure 1.**
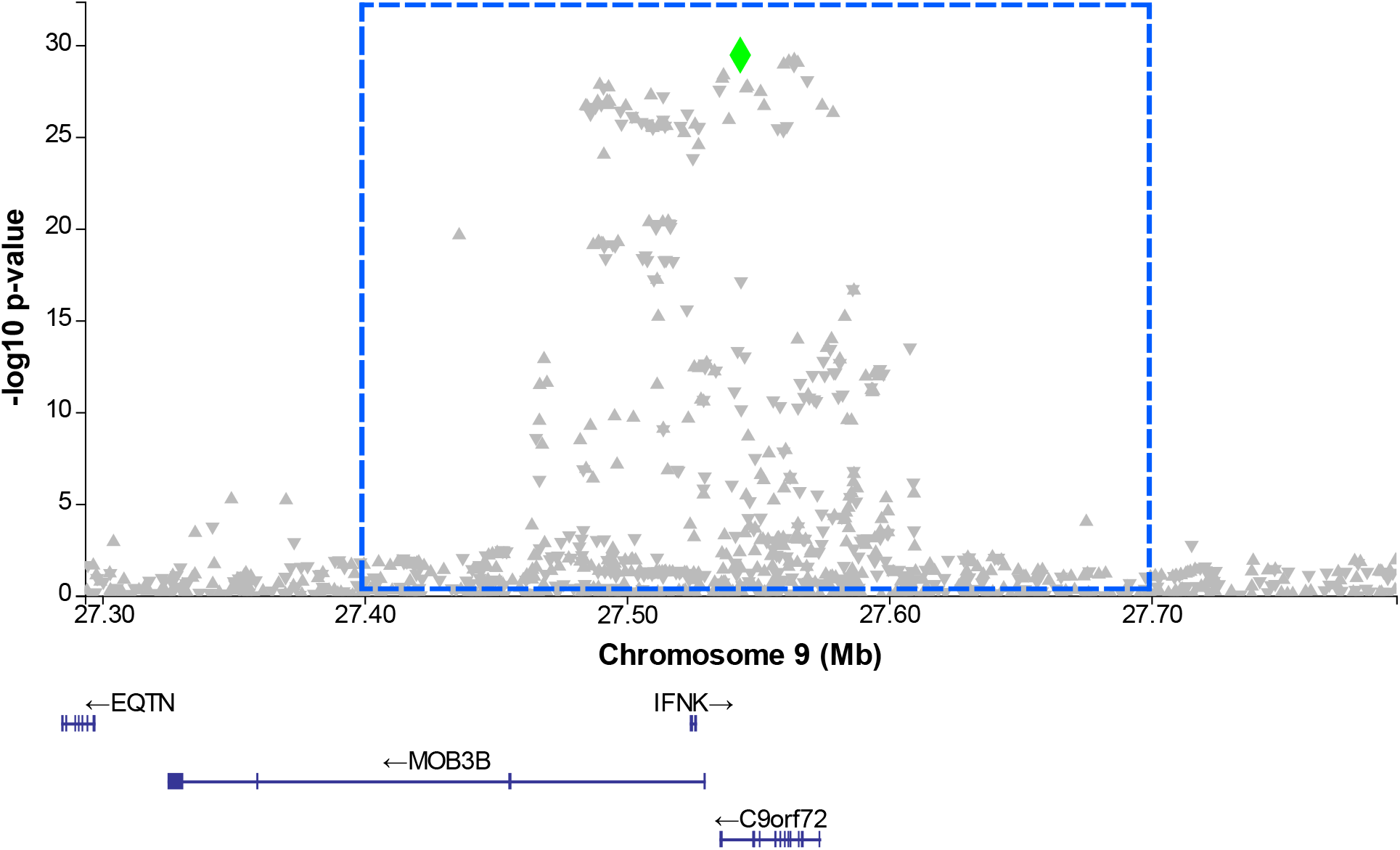
*C9orf72* region of chromosome 9 visualized as a locus zoom plot. Single nucleotide polymorphisms (SNPs) are plotted by genomic position. The y-axis corresponds to -log_10_(p-values) from the ALS genome-wide association study (Nicholas et al. 2018). We considered the *C9orf72* region to span from 27.4 Mb to 27.7 Mb on chromosome 9 as illustrated with the blue dashed box. In an independent sample, our primary polygenic score excluded the *C9orf72* region, and a sensitivity polygenic score included these SNPs. The SNP highlighted by the green diamond (rs3849943, located chr9:27543382) was associated with *C9orf72* repeat status (fisher p-value = 0.00001). Below the plot, positions of *C9orf72* as well as other genes in the region are shown.

Statistical analysis, including summary of case and control populations, regression models, classification attributable fraction calculation, and sensitivity analyses are presented in Supplementary Methods. Gene pathway analysis examining the overall biological functions of polygenic score genes was performed (Supplementary Methods).

Finally, to assess robustness of the polygenic score in the University of Michigan samples, a second Spanish Neurological Consortium cohort that was not part of the ALS GWAS was examined to test for replication (Supplementary Methods).

## Results

### Study Participants

The primary analysis included 442 participants (223 controls and 219 ALS cases) (**Table 1**). Family history of ALS was present in 7.8% of ALS cases and 0% of controls. The *C9orf72* repeat was present in 5.9% of ALS cases and 0% of controls. No age differences occurred between ALS and control participants, although the male participant proportion was higher in the ALS (59.0%) versus control (48%, p-value = 0.027) group. There were 24 participants excluded for missing genetic, demographic, or ALS assessment data (**Supplementary Figure S1B**), who had similar characteristics to the included sample (**Supplementary Table S2**).

**Table 1.** Included study sample characteristics by ALS case and control status for shared ancestry cohort.

| Characteristic | Control<br>N = 223<sup>1</sup> | ALS<br>N = 219<sup>1</sup> | p-value<sup>2</sup> |
| --- | --- | --- | --- |
| **ALS polygenic score with C9orf72 region removed** | -0.08 (-0.73, 0.64) | 0.03 (-0.55, 0.74) | 0.11 |
| **ALS polygenic score with C9orf72 region included** | -0.13 (-0.74, 0.65) | 0.08 (-0.56, 0.75) | 0.080 |
| **C9orf72 Expansion Status** |  |  | <0.001 |
| Negative | 223 (100%) | 206 (94%) |  |
| Positive | 0 (0%) | 13 (5.9%) |  |
| **Family History of ALS** | 0 (0%) | 17 (7.8%) | <0.001 |
| **ALS Onset Segment** |  |  | - |
| Bulbar | 0 (0%) | 59 (27%) |  |
| Cervical | 0 (0%) | 80 (37%) |  |
| Lumbar | 0 (0%) | 73 (33%) |  |
| Respiratory | 0 (0%) | 1 (0.5%) |  |
| Thoracic | 0 (0%) | 4 (1.8%) |  |
| Generalized | 0 (0%) | 2 (0.9%) |  |
| Not applicable | 209 (100%) | 0 (0%) |  |
| **Age (years)** | 65 (58, 71) | 67 (59, 73) | 0.4 |
| **Sex** |  |  | 0.027 |
| Female | 115 (52%) | 90 (41%) |  |
| Male | 108 (48%) | 129 (59%) |  |
| **Self-Reported Race/Ethnicity** |  |  | 0.037 |
| Asian | 2 (0.9%) | 1 (0.5%) |  |
| Black or African American | 11 (5.0%) | 2 (0.9%) |  |
| Hispanic or Latino | 5 (2.3%) | 3 (1.4%) |  |
| White or Caucasian | 203 (92%) | 213 (97%) |  |
| Missing | 2 | 0 |  |
| **Multi-Ancestry Genetic PC1** | 0.0080 (0.0077, 0.0082) | 0.0080 (0.0078, 0.0082) | 0.9 |
| **Multi-Ancestry Genetic PC2** | -0.020 (-0.020, -0.020) | -0.020 (-0.020, -0.019) | 0.6 |
| **Multi-Ancestry Genetic PC3** | -0.0079 (-0.0086, -0.0071) | -0.0080 (-0.0085, -0.0071) | 0.7 |
| **Multi-Ancestry Genetic PC4** | -0.0095 (-0.0101, -0.0089) | -0.0098 (-0.0102, -0.0090) | 0.066 |
| **Multi-Ancestry Genetic PC5** | -0.004 (-0.007, 0.000) | -0.004 (-0.007, -0.001) | 0.8 |

<sup>1</sup>Median (25<sup>th</sup> percentile, 75<sup>th</sup> percentile); n (%)
<sup>2</sup>Wilcoxon rank sum test; Pearson's Chi-squared test; Fisher's exact test
ALS, Amyotrophic lateral sclerosis; N, number; PC, principal component

### Genetic Data Characteristics and Polygenic Score Optimization

SNPs were measured at 1,748,250 positions. SNPs missing genomic location data, with missingness frequency of >1% of samples, with minor allele frequency < 5%, or out of Hardy-Weinberg equilibrium (p-value < 10^−6^) were removed, leaving 601,350 measured autosomal SNPs (**Supplementary Figure S2**). Imputation resulted in 47,109,465 SNPs. Imputed SNPs with an imputation quality R-squared value less than 0.5 and SNPs with a minor allele frequency < 1% were filtered. The final dataset had 8,179,459 imputed SNPs (**Supplementary Figure S3**).

The *C9orf72* region on chromosome 9 spanned from 27.4 Mb to 27.7 Mb. Following pruning, 5 SNPs were present in this region (**Figure 1**). Of these, one SNP rs3849943, located at position 27,543,382, was associated with *C9orf72 expansion* status (fisher p-value = 0.00001). Because our goal was to estimate the cumulative genetic risk for ALS beyond the *C9orf72* expansion, the primary polygenic score excluded this entire region out of caution. We constructed polygenic scores, including SNPs based on their association with ALS in an independent GWAS. Polygenic score performance was highest when constructed using a p-value threshold of approximately 10^−4^, using 275 SNPs (**Supplementary Figure S4**). At this threshold, the incremental R^2^ for the polygenic score was approximately 1.2%.

For sensitivity analyses, we considered a polygenic score using all available SNPs post pruning (n = 254,307 SNPs), with an incremental R^2^ of approximately 0.4%. As another sensitivity analysis, we included SNPs in the *C9orf72* region and observed polygenic score performance was also highest using a p-value threshold of approximately 10^−4^ (n = 280 SNPs) (**Supplementary Figure S5**).

### Associations Between Genetic Predictors and ALS Cases Status

In bivariate analyses, ALS cases had higher mean ALS polygenic scores (average standardized score of 0.03) than controls (average standardized score -0.08) (p-value = 0.11) (**Supplemental Figure S6**). We examined the roles of genetic variables and family history in analyses adjusted for age, sex, and five genetic principal components. In the full study sample (n = 442 participants), a one standard deviation increase in ALS polygenic score was associated with 1.28 times higher odds of ALS (95% CI: 1.04, 1.57) (**Table 2**), after also adjusting for *C9orf72* repeat expansion status and family history of ALS. These findings were consistent when we subset the sample to participants lacking a *C9orf72* repeat or family history of ALS (N = 416 participants). A one standard deviation increase in ALS polygenic score was again associated with 1.28 times higher odds of ALS (95% CI: 1.04, 1.57).

**Table 2.** Regression results in the full sample used in sensitivity analyses (n=223 controls, n=219 ALS cases). Regression results provided are odds ratios and 95% confidence intervals within parentheses for association with ALS status. All Firth penalized logistic regression models were also adjusted for participant age, sex, and 5 genetic ancestry principal components. Polygenic scores for ALS are based on weights in an independent genome-wide association study (Nicholas et al. 2018).^13^

| Variable | C9orf72 region SNPs excluded from polygenic score |  |  | C9orf72 region included |
| --- | --- | --- | --- | --- |
| | Polygenic score<br>($P_{\text{threshold}}=0.0001$,<br>N= 275 SNPs) | Polygenic score<br>($P_{\text{threshold}}=0.0001$,<br>N= 275 SNPs) | Polygenic score<br>($P_{\text{threshold}}=1.0$,<br>N= 254,280 SNPs) | Polygenic score<br>($P_{\text{threshold}}=0.001$,<br>N= 280 SNPs) |
|  | N = 442<br>participants | N = 416<br>participants<br>(without family<br>history and/or<br>C9orf72<br>expansion) | N = 442 participants | N = 442<br>participants |
| Polygenic score<br>(one standard<br>deviation<br>increase) | 1.28 (1.04, 1.57) | 1.28 (1.04, 1.57) | 1.13 (0.77, 1.66) | 1.28 (1.05, 1.58) |
| C9orf72 repeat<br>(positive) | 22.8 (2.8, 2954) | - | 20.7 (2.6, 2674) | 21.4 (2.7, 2775) |
| Family history of<br>ALS (yes) | 33.2 (4.3, 4268) | - | 32.6 (4.3, 4184) | 32.7 (4.2, 4209) |
| Age (10-year<br>increase) | 1.1 (0.91, 1.33) | 1.1 (0.91, 1.33) | 1.09 (0.9, 1.32) | 1.09 (0.9, 1.32) |
| Sex (male) | 1.52 (1.02, 2.27) | 1.52 (1.02, 2.28) | 1.47 (0.99, 2.19) | 1.53 (1.03, 2.29) |

SNP: single nucleotide polymorphism

### ALS Case Classification Performance

Beyond association testing, we were interested in the performance of genetic factors in classifying ALS cases and controls (**Figure 2**). Our base classification model adjusted for sex, age, and five genetic principal components had an area under the curve (AUC) of 0.591. Adding family history of ALS alone to the base model increased AUC to 0.631 and improved classification over the base model (likelihood ratio test p = 0.06). Including *C9orf72* repeat status as a covariate on top of the base model and family history increased the AUC to 0.647 and improved classification (likelihood ratio test p-value < 0.001). Adding the ALS polygenic score following family history and *C9orf72* repeat status further raised AUC to 0.663 and improved classification (likelihood ratio test p-value < 0.001). To assess prediction accuracy, we split datasets into training and testing and performed five-fold cross-validation. These AUC results were 0.539 for the base model, 0.588 adding family history, 0.603 adding *C9orf72* repeat status, and finally 0.620 adding ALS polygenic score (**Supplemental Figure S7**). While the AUCs were attenuated, as a result of the sampling procedure, similar sequential prediction accuracy remained, highlighting the prediction capability.

**Figure 2.**
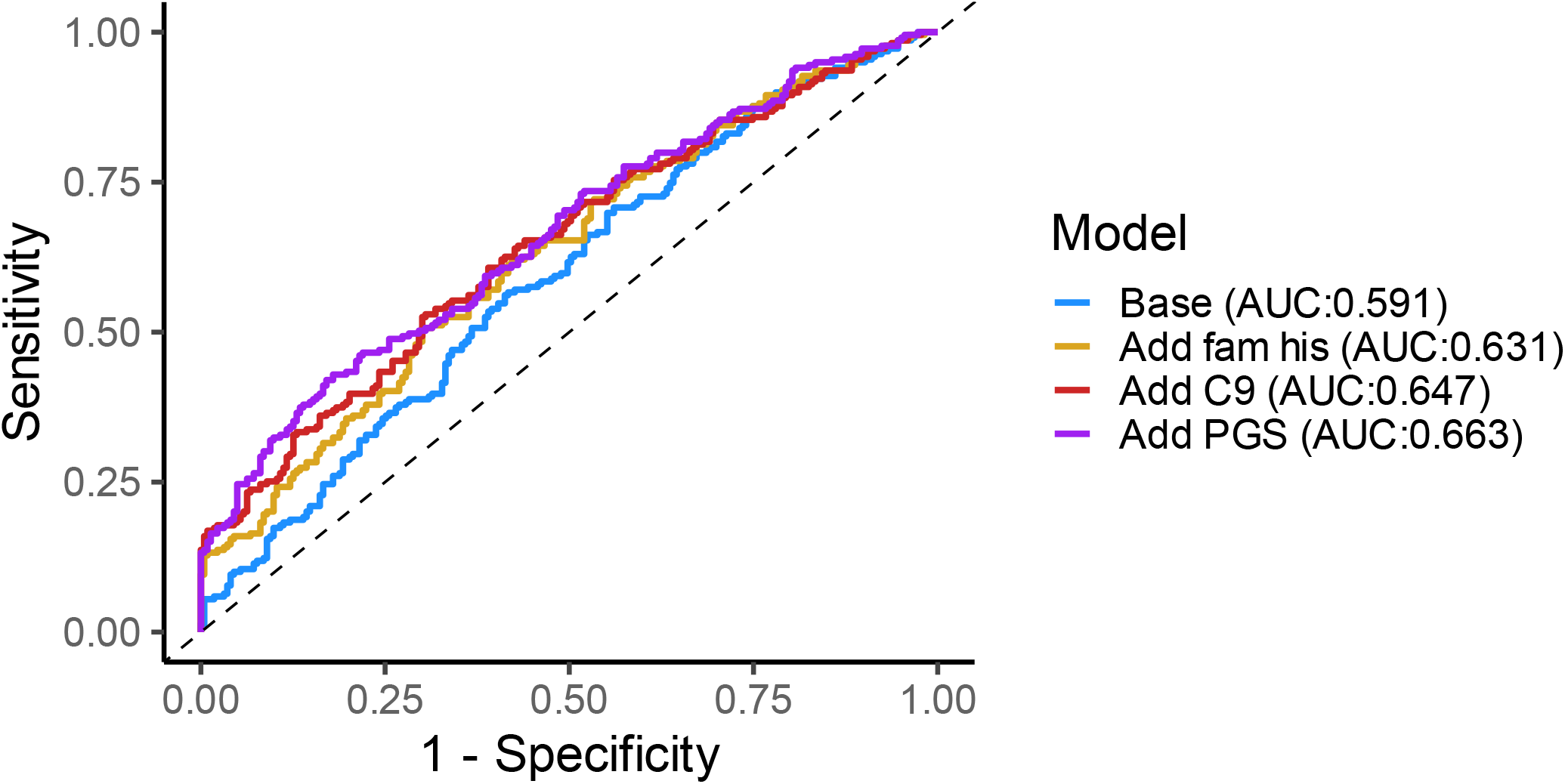
ROC Curve. Base model has sex, age, ancestry principal components. (n = 442) Receiver operating characteristic curve (ROC) for classification of ALS and control participants. The base model includes sex, age, and 5 genetic principal components and has an area under the curve (AUC) of 0.591. Adding family history to the base model increases the AUC to 0.631 (likelihood ratio test p-value = 0.06). Adding *C9orf72* expansion in addition to family history increases the AUC to 0.647 (likelihood ratio test p-value < 0.001). Adding polygenic score (PGS, region around *C9orf72* removed) in addition to family history and *C9orf72* expansion improves the AUC to 0.663 (likelihood ratio test p-value < 0.001).

### Attributable Fraction

To assess the fraction of ALS cases attributable to genetic factors, we compared those in the highest 20^th^ percentile of ALS polygenic score to the rest of the sample. We observed that 4.1% (95% CI: -9.1%, 17.3%) of ALS cases would be prevented if the highest 20th percentile of ALS polygenic score were at the level of the rest of the population. For the *C9orf72* expansion, 6.3% (95% CI: -2.7%, 15.3%) of ALS cases would be avoided if they lacked the expansion.

### Sensitivity Analyses

Sensitivity analysis (Supplementary Results, **Table 2**), including analysis around the *C9orf72* region, and an analysis restricted to European ancestry participants (**Supplementary Table S3, Supplementary Table S3, Supplementary Figure S8**), overall showed findings consistent with the main analysis.

### Gene Pathway Analysis

In the 275 SNP associated genes, included the polygenic score, *richR* identified 65 highly enriched GO biological process terms, including several related to the neuronal system, such as “neuron differentiation”, “generation of neurons”, “neuron projection morphogenesis”, “neurogenesis” and “neuron development” (**Figure 3, Supplementary Table S5**). A total of nine KEGG pathways were significantly enriched at a nominal p-value < 0.05, which included “Glycosphingolipid biosynthesis-ganglio series”, “Fatty acid degradation” and “Pancreatic secretion” (**Figure 4, Supplementary Table S6**).

**Figure 3.**
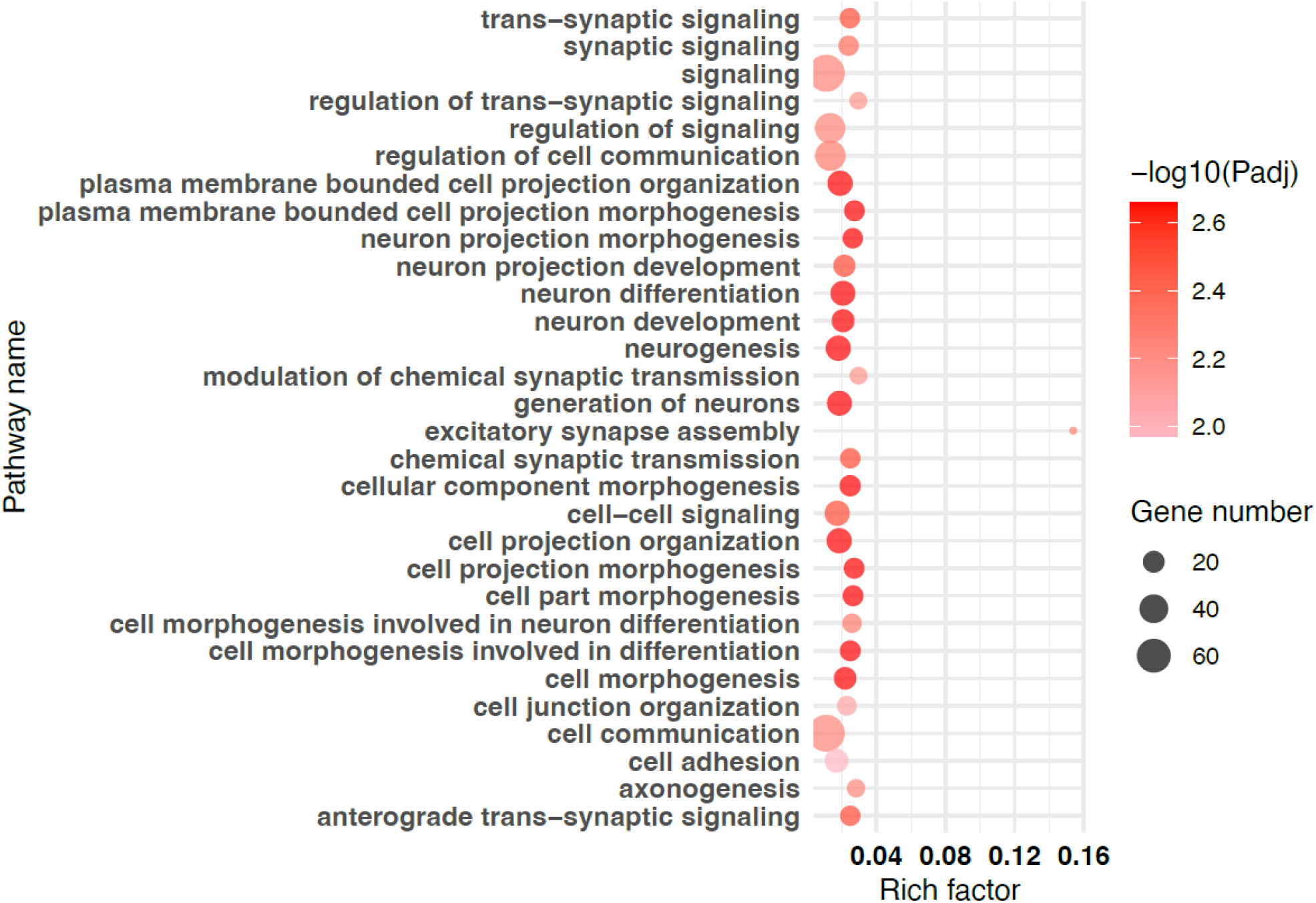
Highly enriched gene ontology (GO) biological processes. The 50 most significantly enriched biological functions using GO are illustrated in dot plots. Rich Factor refers to the proportion of single nucleotide polymorphism (SNP) associated genes belonging to a specific term. The color indicates the level of significance (-log_10_Padj). The numbers correspond to the number of SNP associated genes belong to the term.

**Figure 4.**
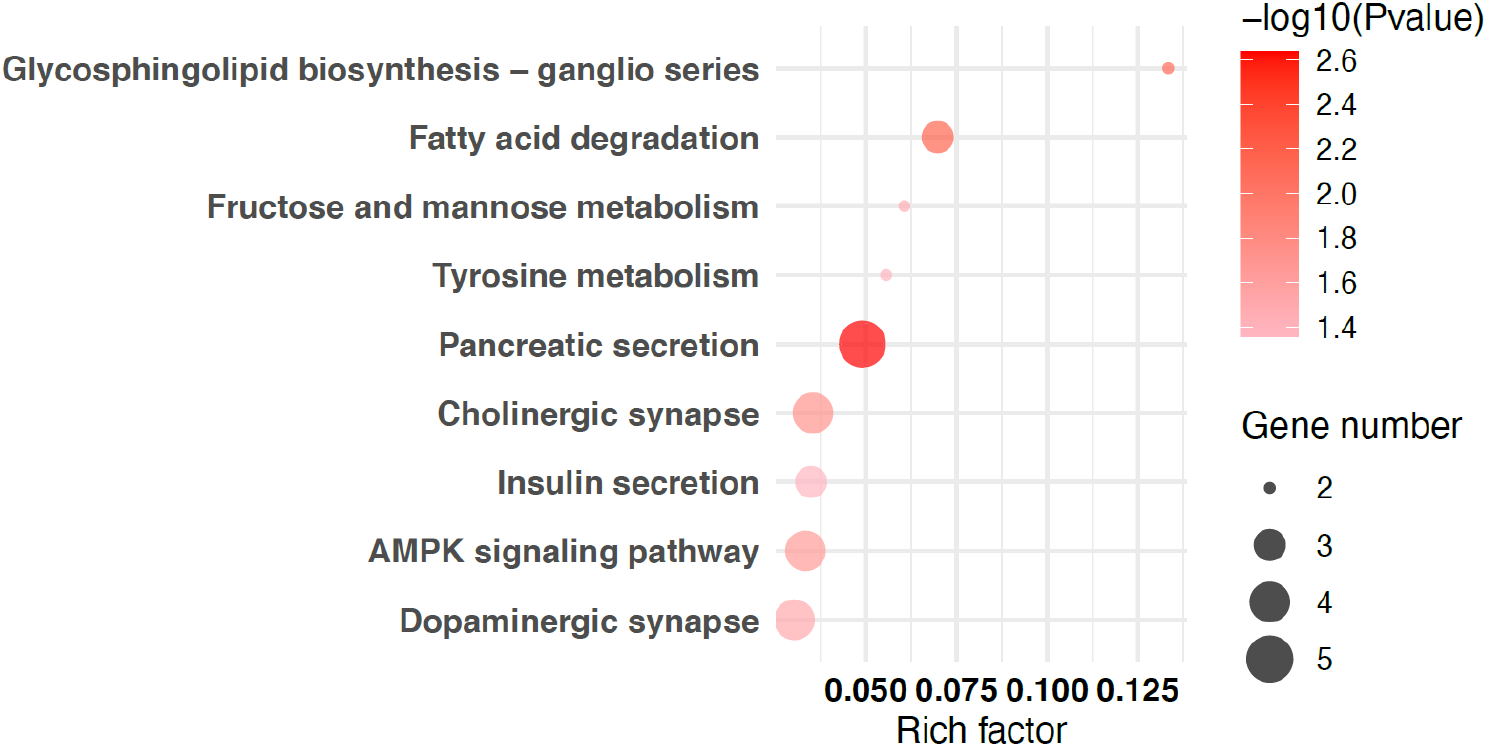
Highly enriched KEGG pathways. The significantly enriched Kyoto Encyclopedia of Genes and Genomes (KEGG) pathways are illustrated in dot plots. Rich Factor refers to the proportion of single nucleotide polymorphism (SNP) associated genes belonging to a specific term. Node size (Gene number) refers to the number of SNP associated genes within each term and node color indicates the level of significance (-log_10_p-value).

### Replication Results

The Spanish cohort had 548 ALS cases and 2,756 controls, after removing 232 participants for missing age or *C9orf72* information. Family history, *C9orf72* expansion, and sex were associated with ALS case status (**Supplementary Table S7**). Due to differences in genotyping arrays and allele frequencies between the Michigan and Spanish cohorts, available SNPs varied between the two cohorts. To harmonize our analyses, we restricted to SNPs available in both cohorts, and the best performance in the Michigan cohort among overlapping SNPs resulted from a polygenic score consisting of 132 SNPs (p-value threshold=5 × 10^−5^). In the Spanish cohort, a one standard deviation increase in the harmonized ALS polygenic score was associated with 1.11 higher odds (95% CI: 1.01, 1.22) of ALS case status (p-value = 0.028), adjusted for sex, age, *C9orf72* expansion, family history, and five genetic principal components. In the Michigan cohort, a one standard deviation increase in the harmonized ALS polygenic score was associated with 1.22 higher odds (95% CI: 1.00, 1.50) of ALS case status (p=0.04) when including all ancestries, mirroring results above with the 275 SNP polygenic score. When limiting to European genetic ancestry in the Michigan cohort, the harmonized 132 SNP polygenic score had a stronger association, where one standard deviation increase in ALS polygenic score was associated with 1.27 higher odds (95% CI: 1.03, 1.57) of ALS case status (p-value = 0.02). Meta-analysis of the Spanish cohort and Michigan cohort (all ancestry) resulted in an estimate of one standard deviation increase in ALS polygenic score being associated with 1.13 higher odds (95% CI: 1.04, 1.23) of ALS case status (p-value = 0.004) (**Supplementary Figure S9**).

## Discussion

ALS risk factors are incompletely understood. Models that predict the steps involved in developing ALS^24^ are necessary to generate ALS risk profiles. Representing this genetic risk^25^ is critical as most individuals with ALS lack a monogenic ALS risk gene. Since genetic risk may be distributed throughout the genome, identifying polygenic risk facilitates an understanding of the multiple ALS pathological pathways. Here we developed a weighted polygenic score using a large ALS-control GWAS.^13^ This score differed significantly in ALS cases versus controls from an independent Michigan cohort. Further, this polygenic score represents important genes and biological functions in the pathophysiology of ALS.

In the current study, the ALS polygenic score with the best model fit and lowest p-value was represented by 275 SNPs when excluding the region around *C9orf72* and 280 SNPs when including the region. We tested other SNP combinations as determined by default PRSice-2 p-value thresholds and a model including all SNPs. In each case, the model with fewer SNPs outperformed the larger models, suggesting that the genetic contributions to ALS are limited to a smaller subset of genes as opposed to a wide-ranging set of genes across more genomic regions. Next, we showed that a standard deviation increase in the ALS polygenic score raised ALS odds by 1.28 times in both models without and with the *C9orf72* region. Interestingly, all risk increased when the *C9orf72* region was included, even after adjusting for the *C9orf72* expansion, suggesting a possible role for alleles around the *C9orf72* region on disease status, even in the absence of the repeat itself. Unsurprisingly, in these models, ALS risk was disproportionate for individuals with a family history or the *C9orf72* expansion. Removing individuals with an ALS family history or a *C9orf72* expansion did not change the impact of the polygenic score on ALS risk, meaning the polygenic score itself plays an essential role on the overall ALS risk profile. Additionally, findings persisted when restricting to a European genetic ancestry population.

Polygenic scores summarize the combined effects that common and low-frequency alleles have on disease risk, thereby summarizing the genetic architecture of that disease.^26^ Multiple medical specialties utilize polygenic scores to explain risks such as cardiovascular disease, cancers, neurodegenerative diseases,^19 26^ and other phenotypic traits.^10^ While polygenic scores are gaining traction for ALS,^27^ few studies propose an ALS-specific polygenic score that can stratify populations at risk for ALS.

In contrast to our methods, McCann and colleagues leveraged a list of 853 genetic variants with a changed amino acid sequence from a comprehensive literature search.^28^ After screening the population, 43 genetic variants from 18 genes were retained in the model, affecting 35.4% of their ALS population. However, the authors did not further develop polygenic scores.^28^ Wainberg et al. identified individuals in the Arivale Scientific Wellness cohort at elevated genetic risk for ALS using polygenic risk scores developed through literature and sought linkages to proteomics, metabolomics, and other clinical laboratory information. This group found that increased Ω-3 and decreased Ω-6 fatty acid levels and higher IL-13 levels correlated with ALS genetic risk.^27^ Based on KEGG analysis of the polygenic score developed herein, we found enrichment of the fatty acid degradation pathway.^29^

Placek and colleagues used sparse canonical correlation analysis to identify a polygenic score of cognitive dysfunction in an ALS population.^30^ Like our methods, the authors focused on SNPs achieving genome-wide significance in the Nicolas study^13^ and with risk loci in ALS and frontotemporal dementia. Of the 45 SNPs used in their models, 27 were associated with cognitive performance in their ALS population, involving the genes *MOBP, NSF, ATXN3, ERGIC1*, and *UNC13A*. Our polygenic score also included SNPs in MOBP, ATXN3, and UNC13A, thereby supporting its validity. Additional uses of polygenic scores in ALS include examining polygenic traits for other diseases that overlap with ALS.^10 31^ Although this was not our approach, such studies have yielded linkages between ALS and traits of schizophrenia, cognitive performance, and educational attainment.^10 31^

Polygenic scores have shown utility in other neurodegenerative conditions, such as Alzheimer’s disease, to find those at high and low genetic risk.^32^ For example, a polygenic score derived from the International Genomics of Alzheimer’s Disease Project GWAS showed it could predict participants that would transition from mild cognitive impairment to late-onset Alzheimer’s disease.^33^ A similar approach using a polygenic score created from an Alzheimer’s cohort GWAS dataset associated with incident dementia in a large Swedish birth cohort.^34^

Our disease classification model further supports the utility of our polygenic score. Our polygenic score improved model performance, even one that included the most prevalent ALS risk gene, the *C9orf72* expansion. In Alzheimer’s, similar findings are noted, where a polygenic risk score was able to classify Alzheimer’s cases versus controls with an AUC of 0.83, even when excluding *APOE4* carriers.^35^ This indicates that these genetic models are beneficial in case classification, even when considering strong genetic risks, which superimpose on polygenic risk. Another analysis similarly showed that polygenic scores in Alzheimer’s disease could classify patients accurately and that the prediction improved when incorporating additional variables such as sex and age.^36^

Since polygenic scores often overlap in persons with and without a disease of interest, focusing on patients with polygenic scores in distribution tails may offer better predictive power.^37^ Thus, to add further perspective to this polygenic risk, we showed that 4.1% of ALS cases could be avoided for individuals with the highest 20% of polygenic score if an intervention were possible. While this population attributable risk approach considers the fraction of disease caused by exposure, this idea can also be applied to genetic data.^38 39^ For example, a study of polygenic scores in cutaneous squamous cell carcinoma showed that removing all risk alleles from a population would decrease the risk of this cancer by 62%.^40^ The authors argue that identifying those at the highest genetic risk could inform programs for skin cancer screening, with the caveat that interactions of SNPs with environmental factors^41^ are not included in the model. A parallel approach is also proposed for breast cancer to help identify populations that would benefit from targeted risk reduction strategies.^42^ A similar analysis has shown changes in the prevalence of type 2 diabetes, breast cancer, hypertension, and myocardial infarctions, if a proportion of polygenic risk is removed or enhanced in the population.^43^ Currently, there is no biomarker or tool that can definitively predict who will develop ALS later in life. Therefore, even if the polygenic score can only explain a small number of individuals at risk, it could be an important screening method for risk reduction.

Replication of these findings is important to determine the generalizability of the results. We used genotype and ALS phenotype data from an independent Spanish cohort as a replication cohort. Although the SNPs included in the polygenic score were adjusted due to the available overlap of SNPs in both datasets, there was consistency in the magnitude of the polygenic score effect, further providing support for our proposed polygenic score. Replication of polygenic scores is critical to ascertain that the methods and population background used to develop the score is generalizable.^37^ Further, replication cohorts can determine which risk variants are applicable across diverse populations.^44^ Replicating polygenic scores in ALS remains important, although this requires large numbers of samples from participants not included in GWAS used to derive SNP weights.

We next queried how this set of SNPs impacts disease pathobiology. Through gene enrichment and pathway analysis, we showed that this polygenic score selects multiple pathways relevant to ALS biology, including synaptic signaling, regulation of protein metabolic process, neuron projection, and axon guidance. Using KEGG pathways, we also identified important ALS biological functions, including glycosphingolipid biosynthesis and fatty acid degradation.^16 45^ Saez-Atienzar et al. used a cohort of 78,500 individuals to develop a polygenic score for biological pathways and cell types to determine involvement in ALS.^46^ Significant pathways included those involved in neuronal development and differentiation with an emphasis on the cytoskeleton. Of these pathways, the cytoskeleton pathway was significant for individuals both with and without the *C9orf72* repeat expansion, whereas the autophagosome pathway was only significant for *C9orf72* carriers. Overlapping enriched GO pathways in our polygenic score with those of Saez-Atienzar et al. included neuron projection morphogenesis, cell morphogenesis involved in differentiation, neuron development, cellular component morphogenesis, cell development, and cell projection organization. The overall overlap shows that these two different methods for developing a polygenic score selects similar pathways. Other studies of gene expression in ALS have also identified dysregulated metabolic pathways and cytoskeletal pathways.^47^

This study has limitations. Due to cost and a research interest in common genetic variants, we performed genome-wide genotyping instead of whole genome sequencing. While whole genome sequencing would allow us to better account for genetic background, the method we used are validated across many studies. In addition, the study population size is small compared to the number of individuals impacted by ALS. However, the sample size here was limited to participants not included in prior GWASs and is thus a strength. This is important since developing polygenic scores from participants that are already in the reference GWAS may lead to biased results. Also, since we did utilize a lower-cost genotyping strategy imputed to maximize overlap with the ALS GWAS used for weights, these methods could be beneficial for population screening where the cost of whole genome sequencing is not economically feasible.

## Conclusion

In conclusion, we find that a polygenic score for ALS can account for cumulative genetic risk in the population and reflect cellular processes that are relevant to ALS. If further validated, this polygenic score can be a valuable tool for ALS risk models and the design of ALS prevention studies.

## Data Availability

Data may be shared by qualified investigators by reasonable request to the corresponding author. A data request proposal is reviewed and approved by a review panel, and a signed data-sharing agreement will then be approved. Code to perform preprocessing and analyses is available (https://github.com/bakulskilab).

## Acknowledgments

We thank Crystal Pacut, Caroline Piecuch, and Stacey Sakowski, PhD for study support, Masha Savelieff, PhD and Emily Koubek, PhD, for editorial assistance, and the following members of the Spanish Neurological Consortium for study assistance: Mónica Povedano Panadés, Antonio Guerrero-Sola, Tania García-Sobrino, Marilina Puente Hernández, María Jesús Sobrido-Gómez, Ivonne Jericó-Pascual, Carmen Paradas López, Adriano Jimenez-Escrig, Mario Ezquerra, and Ana Gorostidi Pagola. This study utilized the high-performance computational capabilities of the Biowulf Linux cluster at the NIH.

## Authors’ contributions

Substantial contributions to the conception or design of the work: KMB, SS-A, BJT, ELF, and SAG. Acquisition, analysis, or interpretation of data for the work: JFD, KMB, KG, JH, LZ, SS-A, ARS, RC, AG-R, RR-G, JFV-C, RFS, SB-C, PG-C, MTP, PM, JP-T, FC, MM-G, JR, DB-H, LG-D, JIC, PP, BJT, ELF, and SAG. Drafting the work or revising it critically for important intellectual content: JFD, KMB, KG, JH, LZ, SS-A, ARS, RC, AG-R, RR-G, JFV-C, RFS, SB-C, PG-C, MTP, PM, JP-T, FC, MM-G, JR, DB-H, LG-D, JIC, PP, BJT, ELF, and SAG.

