## Supplemental Material for "Cumulative genetic risk and *C9orf72* repeat status independently associate with ALS status in two case-control studies"

### Supplementary information

This document contains Supplemental Methods, Supplemental Results, 9 Supplemental Figures, and 7 Supplemental Tables.

### Supplementary Methods

*Michigan Study Participants and Sample Collection*

All patients seen at the University of Michigan Pranger ALS Clinic are invited to participate, although the present case/control analysis is limited to those with ALS, thereby excluding participants with other forms of motor neuron disease. Healthy controls, without a personal or family history of a neurodegenerative disease in a first- or second-degree family member, are identified using a recruitment database available through the Michigan Institute for Clinical & Health Research and through population outreach via random address mailings. Participant demographics including sex (male, female), race/ethnicity (White or Caucasian, Black or African American, or Asian and Hispanic or Latino), and age (years) were obtained at the time of study enrollment. ALS diagnoses were confirmed by an ALS neurologist (S.A.G., E.L.F.), who also recorded onset age (years), diagnosis age (years), onset segment (bulbar, cervical, lumbar, respiratory, thoracic), and presence of an ALS family history (yes or no) in the medical record. A family history of ALS in a first- or second-degree relative is considered positive. All participants provide venous blood, collected in an EDTA tube and frozen at -80 °C for later DNA extraction. The study is approved by the University of Michigan Institutional Review Board (HUM28826) and all participants provide written informed consent.

*Spanish Neurological Consortium*

The replication cohort used here includes 3536 individuals (648 participants with ALS and 2888 neurologically healthy controls). Most controls (2823/2888) were recruited from several centers across Spain, as reported in a previous study.[1] For the Spanish Neurological Consortium samples, analysis of samples was approved by the institutional review board of the National Institute on Aging (protocol number 03-AG-N329). Written consent was obtained from all individuals enrolled. The participants were recruited through several centers in Spain, where ethics approval of samples was obtained from each participating institution: Universidad de Sevilla, University Hospital Mutua de Terrassa, Hospital Universitario Central de Asturias, Universidad de Granada, Instituto de Investigación Sanitaria Biodonostia, Universidad de Murcia, Hospital Clinic de Barcelona, Hospital General de Segovia, Hospital de la Santa Creu i Sant Pau, Universitat Autònoma de Barcelona and Centro de Investigación Biomédica en Red en Enfermedades Neurodegenerativas (CIBERNED), Hospital Gregorio Marañon, Instituto de Investigación Biosanitaria de Granada, Instituto de Investigación Sanitaria Fundación Jiménez Díaz, Hospital Universitario Virgen de la Victoria, Hospital Universitario Marqués de Valdecilla-IDIVAL, Instituto de Investigación Sanitaria Biodonostia, Institut de Recerca Sant Joan de Déu, Hospital Universitario Ramón y Cajal, Hospital Universitario y Politécnico La Fe, Hospital Clinic Barcelona, Hospital Universitario Infanta Sofía, Hospital Sant Pau, Hospital Universitario Virgen de la Victoria, Hospital Universitario Donostia, Instituto de Investigación Sanitaria Biodonostia, Hospital Universitario Marqués de Valdecilla-IDIVAL, Hospital Universitario Marqués de Valdecilla-IDIVAL, Hospital General de Segovia, and Hospital Universitario Fundación Alcorcón.

The remaining control samples (65/2888) and the ALS samples (648) were recruited as a part of the ALS Genetic Spanish Consortium (ALSGESCO).[2] Spanish Neurological Consortium ALS samples were obtained from the ALS Genetic Spanish Consortium. Written consent was obtained from all individuals enrolled. The participants of the ALS Genetic Spanish Consortium were recruited through several motor neuron disease centers in Spain, where ethics approval of samples was obtained from each participating institution: Instituto de Investigación Biomédica Hospital 12 de Octubre, Hospital Universitario Gregorio Marañón, Hospital Carlos III, Hospital Universitario Basurto, Hospital de la Fe, Hospital Clínico Universitario "San Carlos", Institut de Biomedicina de Valencia-CSIC, Hospital Universitario Donostia, Hospital Universitario Virgen del Rocío, Hospital Clinic of Barcelona, Hospital Clínico Universitario "Lozano Blesa", Hospital

Miguel Servet, Hospital del Mar, Hospital General Yagüe, Hospital Nuestra Señora de Valme, Hospital Virgen del Camino, Hospital de Getafe, Hospital General Universitario de Alicante, Hospital Severo Ochoa, Hospital de la Santa Creu i Sant Pau. Participants were diagnosed according to the revised El Escorial Criteria. They were diagnosed and followed by neurologists specialized at Motor Neuron Diseases Units included in ALSGESCO. The study of *C9orf72* hexanucleotide expansion were carried out by triple PCR methodology as described before in the laboratories included in ALSGESCO.[2] Of these samples, 31 ALS cases were missing *C9orf72* expansion status and were excluded, 69 cases were missing age and were excluded, and 132 controls were missing age and were excluded.

*DNA Analysis*
DNA was extracted using the QIAamp DNA Kit (Qiagen, Venlo, Netherlands). Genome-wide genotypes at 1,748,250 positions were measured for 512 samples using the Infinium Multi-Ethnic Global-8 v1.0 array kit (Illumina, San Diego, CA) by the University of Michigan Advanced Genomics Core (**Supplementary Figure S1A**). All available clinical samples, including intentional duplicates (n=6) and non-ALS diseased samples (6 primary lateral sclerosis, 12 other motor neuron disease), were included at this step to improve imputation quality. DNA samples were also analyzed for the presence of the *C9orf72* repeat expansion per published protocols.[3]

The PLINK (version 1.9) program performed genetic microarray data quality control checks.[4] Participants and single nucleotide polymorphisms (SNPs) were filtered using recommended thresholds.[5, 6] Participants were excluded for missing data at greater than 1% of SNPs, discrepancies between genetic sex and predicted sex, and heterozygosity greater than three standard deviations from the mean. For intentional technical duplicate samples and unintentional related samples, the sample in each pair with the highest missingness was excluded. Participant inclusion based on genetic data quality control was visualized using a flow diagram and 488 unique ALS and control participants met study inclusion criteria and genetic quality filtering (**Supplementary Figure S1B**).

SNPs were excluded for missing genomic location data or missingness frequency in over 1% of samples. SNPs from autosomal chromosomes and the pseudo-autosomal region of the sex chromosomes were handled separately from the non-autosomal regions of the sex chromosomes. Autosomal and pseudo-autosomal region SNPs were further excluded for minor allele frequency less than 5% or for violating Hardy-Weinberg equilibrium (p-value < $10^{-6}$). Hardy-Weinberg equilibrium was not evaluated in non-autosomal SNPs for males. SNP exclusion was described using a flow diagram and 610,350 measured autosomal SNPs remained (**Supplementary Figure S2).**

Population stratification by genetic ancestry can lead to confounding in genetic analyses.[7] Principal components were computed to identify genetic ancestry groups in our sample merged with the 1000 genomes version 5[8] reference panel. Individuals of all genetic ancestries were included in the main analysis, which adjusted for the first five multi-ancestry principal components. A sensitivity analysis was performed limited to those participants with European ancestry by only taking those clustered with known European ancestry 1000 genomes samples (principal component 1 < 0.02, principal component 2 < 0.08). Principal components were recomputed within the European ancestry sample and used the first five principal components as adjustment covariates.

To harmonize with the ALS GWAS,[9] measured and cleaned genetic data were imputed with 1000 genomes version 5[8] using the Minimac4 program.[10] Following imputation, SNPs were filtered out if they had an imputation quality $R^2 > 0.5$ or a minor allele frequency < 1% in the study sample and described using a flow diagram (**Supplementary Figure S3**).

*Polygenic Score Development*
The imputed and cleaned SNP data created a polygenic score for ALS risk in the study sample. ALS risk weights for the SNPs came from a GWAS of 20,806 ALS cases and 59,804 controls.[9] Eligible SNPs were filtered from those present in the ALS GWAS and this study's cleaned and imputed data. The PRSice 2.0 program was used to create polygenic scores.[11] Default program parameters for pruning and clumping (250kb window, $R^2$ threshold 0.1) were used to account for linkage disequilibrium. Polygenic scores were created as the sum of the weighted number of variant alleles per individual. SNPs were included in the polygenic scores at a series of p-value thresholds from the parent GWAS ranging from low p-values (only most significant SNPs) to a 1.0 p-value threshold (using all SNPs). The polygenic score with the highest $R^2$ in relation to ALS case-control status was selected for our primary analyses.

To assess the cumulative genetic risk for ALS contributed by SNPs located beyond the *C9orf72* genomic region, we excluded chromosome 9 SNPs between 27,400,000 and 27,700,000 base pair positions when generating the primary polygenic score. We recalculated the polygenic score as a sensitivity analysis, allowing SNPs in this *C9orf72* genomic region to be included. To visualize SNPs in the *C9orf72* region in more detail, we used a locus zoom plot[12] (**Figure 1**). We tested any SNPs in this region for correlation with *C9orf72* expansion status using Fisher's exact test.

*Statistical Analyses*
Statistical analyses were performed in R statistical software (version 4.1). Samples were excluded from statistical analysis if they were duplicates or if they were from non-ALS or control participants (n=17 non_ALS cases, n=5 at-risk controls). Next, participants were excluded (n=24) for missing data key covariates (sex, family history, age, *C9orf72* expansion status). The primary analysis included all genetic ancestries. The distributions of continuous covariates were described using mean and standard deviation and the distributions of categorical covariates were described using number and percent of the sample. We provided distributions of covariates separately among the included and excluded samples. Among the included sample, distributions of covariates separately by ALS and controls are provided. Wilcoxon rank-sum test for continuous covariates and chi-square or Fisher's exact test for categorical covariates tested for differences in the distributions of covariates between ALS and control participants.

All regression models were adjusted for sex, age, family history of ALS, and five genetic principal components. The first analysis used multivariable logistic regression to test for an association between ALS and control status with ALS polygenic score. The second model tested for an association with *C9orf72* expansion status. The third model tested both genetic components (ALS polygenic score and *C9orf72* expansion status) as predictors. Since family history and *C9orf72* expansion status had zero cell counts with no controls, Firth penalized likelihood regression was used to avoid unstable effect estimates. We reported odds ratios, 95% confidence intervals, and p-values for predictors of interest, and we reported adjusted $R^2$ for model fit.

To quantify the classification of the genetic predictors on ALS and control status, we used receiver operating characteristic curves to estimate the areas under the curve (AUC) and C-statistics. Our base model was adjusted for sex, age, and five genetic principal components. We compared the AUC from the three genetic models, sequentially adding family history, *C9orf72* expansion, and the ALS polygenic score to the base model. We used a likelihood ratio test to assess whether adding ALS genetic risk covariates improved classification ability over the base model. Five-fold cross-validation was used to compute predicted probabilities and then calculate the AUC to prevent model overfitting.

The attributable fraction was calculated to determine the proportion of the population burden due to ALS that would be avoided without the cumulative genetic risk described in the polygenic score. Those in the highest $20^{th}$ percentile of ALS polygenic score were first compared to the rest in the lowest $80^{th}$ percentile of ALS polygenic score. The population attributable fractions and confidence intervals were calculated using the case-control option in the AF package.[13]

*Sensitivity Analyses*
We tested a multiplicative interaction term between polygenic score and *C9orf72* expansion in the multivariable logistic regression model to assess potential interaction between the polygenic score and *C9orf72* expansion status. To test the impact of retaining SNPs in the *C9orf72* region, a version of the polygenic score was generated containing SNPs in the *C9orf72* region. We repeated the multivariable logistic regression analyses in the full sample using the polygenic score containing the *C9orf72* region. A further sensitivity analysis excluded those with an ALS family history and/or *C9orf72* expansion and conducted the multivariable logistic regression, adjusting for sex, age, and five genetic principal components.

To account for potential population stratification, we subset the study sample to those with European genetic ancestry and repeated the analyses. In the European ancestry subsample, we conducted bivariate descriptive analyses, performed multivariable logistic regression (adjusting for the sex, age, family history of ALS, and five genetic principal components calculated within the European ancestry sample), and tested the classification of ALS cases and controls.

*Gene Pathway Analysis*
To examine the overall biological functions of genes involved in the polygenic score, we annotated the 275 polygenic score SNPs to their nearest gene based on the human hg19 reference genome annotation. Then the functional enrichment analysis for Gene Ontology (GO biological process and Kyoto Encyclopedia of Genes and Genomes (KEGG) pathway was performed by using our in-house *richR* analysis package (https://github.com/hurlab/richR). We considered significance cutoffs as Benjamini-Hochberg adjusted p-value < 0.05 for GO and nominal p-value < 0.05 for KEGG.

*Replication Testing*
A second independent cohort, the Spanish Neurological Consortium, that was not part of the ALS GWAS was examined to test for replication. Samples were analyzed on the NeuroChip Array (v.1.0 or v.1.1; Illumina, San Diego, CA) and processing followed published methods.[1] The Spanish cohort did not have measures for all the SNPs present in the Michigan cohort's polygenic score. Thus, overlapping SNPs in both datasets were selected to harmonize between the cohorts. Using the reduced SNP list, the same procedure described above was repeated in the Michigan cohort to optimize a polygenic score. Selected SNPs were then used to create a polygenic score in the Spanish cohort. Separate logistic regression models fit each cohort using the harmonized polygenic score. Regression models were adjusted for sex, age, family history of ALS, *C9orf72* expansion status, and five genetic principal components. Results from the two cohorts were then meta-analyzed together.

### Supplementary Results

*Sensitivity Analyses*
To assess potential effect modification between the polygenic score and *C9orf72* expansion status, we tested a multiplicative interaction term, which was not significant (p-value = 0.60). To assess the selection of SNPs for inclusion in the polygenic score, we also used an ALS polygenic score constructed with all SNPs (n = 254,280 SNPs), regardless of the p-value of association with ALS, which attenuated the association between the polygenic score and ALS status. A one standard deviation increase in the ALS polygenic score was associated with 1.13 times higher odds of ALS (95% CI: 0.77, 1.66), suggesting that adding all SNPs introduces noise into the polygenic score (**Table 2**). Further, we observed a slightly stronger association when considering the polygenic score (p-value threshold = 10-4), which included SNPs in the region around C9orf72. A standard deviation increase in ALS polygenic score was associated with 1.28 times higher odds of ALS (95% CI: 1.05, 1.58) (**Table 2**).

Genetic analyses can be subject to population stratification related to genetic ancestries. As a sensitivity analysis, we limited our analysis to 409 participants of European ancestry (202 controls and 207 ALS cases) (**Supplementary Table S3**). In the European genetic ancestry subsample, the ALS polygenic score also associated with ALS case status. A one standard deviation increase in polygenic score associated with 1.27 times higher odds of ALS (95% CI: 1.03, 1.57) (**Supplementary Table S4**). As with the full sample, in the European ancestry only sample, the ALS polygenic score containing the *C9orf72* region was more strongly associated, where one standard deviation increase in polygenic score associated with 1.28 times higher odds of ALS (95% CI: 1.05, 1.58).

In the European ancestry only subsample, ALS case-control classification was similarly improved by adding ALS polygenic score to a base model of sex, age, and five genetic principal components. AUC was 0.569 in the base model while adding family history of ALS alone increased AUC to 0.611 (likelihood ratio test p-value = 0.42). Adding *C9orf72* repeat status increased AUC to 0.630 (likelihood ratio test p-value = 0.01), whereas including the ALS polygenic score (*C9orf72* region removed) further enhanced AUC to 0.647 (likelihood ratio test p-value < 0.001) (**Supplementary Figure S8**).

**Tables**

*Supplementary Table S1. Single nucleotide polymorphisms (SNPs) included in the polygenic score.*

For each SNP included in the polygenic score, this table contains the identifier, chromosome, position, weight, and p-value. The SNPs are from an ALS genome-wide association study of 20,806 ALS cases and 59,804 controls[9] that was used as weights in our polygenic score. This table is provided as a separate file labeled “Supplemental Table 1 – Polygenic Score SNPs.csv” and is consistent with Polygenic Score Reporting Standards.

*Supplementary Table S2. Comparison of characteristics between included analytic and excluded sample.*

| Characteristic | Analytic, N = 442[1] | Excluded, N = 24[1] | P-value[2] |
|---|---|---|---|
| **ALS Case/Control Status** | | | >0.9 |
| Control | 223 (50%) | 12 (50%) | |
| Case | 219 (50%) | 12 (50%) | |
| **ALS polygenic score with C9 Region Removed** | -0.02 (-0.66, 0.70) | -0.36 (-0.69, 0.43) | 0.4 |
| **ALS polygenic score with C9 Region Included** | -0.04 (-0.63, 0.65) | -0.45 (-0.76, 0.25) | 0.3 |
| ***C9orf72* Expansion Status** | | | >0.9 |
| Negative | 429 (97%) | 12 (100%) | |
| Positive | 13 (2.9%) | 0 (0%) | |
| Missing | 0 | 12 | |
| **Family History of ALS** | | | 0.016 |
| Yes | 17 (3.8%) | 3 (23%) | |
| No | 425 (96.2%) | 10 (77%) | |
| Missing | 0 | 11 | |
| **ALS Onset Segment** | | | 0.4 |
| Bulbar | 59 (27%) | 1 (8.3%) | |
| Cervical | 80 (37%) | 4 (33%) | |
| Lumbar | 73 (33%) | 7 (58%) | |
| Respiratory | 1 (0.5%) | 0 (0%) | |
| Thoracic | 4 (1.8%) | 0 (0%) | |
| Generalized | 2 (0.9%) | 0 (0%) | |
| Not applicable | 223 | 12 | |
| **Age (years)** | 66 (58, 72) | 64 (54, 74) | 0.4 |
| Missing | 0 | 1 | |
| **Sex** | | | 0.4 |
| Female | 205 (46%) | 9 (38%) | |
| Male | 237 (54%) | 15 (62%) | |
| **Self-Reported Race/Ethnicity** | | | 0.6 |
| Asian | 3 (0.7%) | 0 (0%) | |
| Black or African American | 13 (3.0%) | 0 (0%) | |
| Hispanic or Latino | 8 (1.8%) | 1 (4.2%) | |
| White or Caucasian | 416 (95%) | 23 (96%) | |
| Missing | 2 | 0 | |

[1]n (%); Median ($25^{th}$ percentile, $75^{th}$ percentile)
[2]Pearson's Chi-squared test; Wilcoxon rank sum test; Fisher's exact test
ALS, Amyotrophic lateral sclerosis; N, number

*Supplementary Table S3. Sample characteristics of the European genetic ancestry only sample used in sensitivity analyses.*

| Characteristic | Control N = 202[1] | ALS Case N = 207[1] | P-value[2] |
|---|---|---|---|
| **ALS polygenic score with *C9orf72* region removed** | -0.11 (-0.84, 0.58) | 0.00 (-0.57, 0.67) | 0.068 |
| **ALS polygenic score with *C9orf72* region included** | -0.16 (-0.90, 0.58) | 0.06 (-0.57, 0.64) | 0.046 |
| ***C9orf72* repeat status** | | | <0.001 |
| Negative | 202 (100%) | 194 (94%) | |
| Positive | 0 (0%) | 13 (6.3%) | |
| **Family History of ALS** | | | <0.001 |
| No | 189 (100%) | 192 (92.8%) | |
| Yes | 0 (0%) | 15 (7.2%) | |
| **ALS Onset Segment** | | | - |
| Bulbar | 0 (0%) | 56 (27%) | |
| Cervical | 0 (0%) | 74 (36%) | |
| Lumbar | 0 (0%) | 70 (34%) | |
| Respiratory | 0 (0%) | 1 (0.5%) | |
| Thoracic | 0 (0%) | 4 (1.9%) | |
| Cannot be determined | 0 (0%) | 2 (1.0%) | |
| Not applicable | 189 (100%) | 0 (0%) | |
| **Age** | 66 (59, 72) | 67 (59, 73) | 0.6 |
| **Sex** | | | 0.034 |
| Female | 106 (52%) | 87 (42%) | |
| Male | 96 (48%) | 120 (58%) | |
| **Within Euro PC1** | 0.004 (-0.001, 0.008) | 0.004 (0.000, 0.007) | >0.9 |
| **Within Euro PC2** | -0.020 (-0.028, -0.010) | -0.019 (-0.027, -0.009) | 0.7 |
| **Within Euro PC3** | 0.01 (-0.01, 0.03) | 0.01 (-0.01, 0.02) | 0.4 |
| **Within Euro PC4** | -0.015 (-0.030, 0.000) | -0.016 (-0.028, 0.001) | 0.8 |
| **Within Euro PC5** | -0.006 (-0.027, 0.018) | -0.009 (-0.025, 0.016) | 0.7 |

[1]Median (IQR); n (%)

[2]Wilcoxon rank sum test; Pearson's Chi-squared test; Fisher's exact test

PC, principal component

*Supplementary Table S4. Regression results in the European genetic ancestry sample used in sensitivity analyses (n = 187 controls, n = 216 ALS cases).*

Regression results provided as odds ratios (95% confidence intervals) associated with ALS status. All Firth penalized logistic regression models were also adjusted for participant age, sex, and 5 genetic ancestry principal components. Polygenic scores for ALS are based on weights in an independent genome-wide association study (Nicholas et al. 2018).[9]

| Variable | **Polygenic score excluding *C9orf72* region SNPs**<br>N = 409 participants<br>N = 275 SNPs in polygenic score | **Polygenic score including *C9orf72* region SNPs**<br>N = 409 participants<br>N = 280 SNPs in polygenic score | **Polygenic score excluding *C9orf72* region SNPs**<br>N = 385 participants<br>N = 275 SNPs in polygenic score | **Polygenic score including all SNPs ($P_{threshold}$=1.0)**<br>N = 409 participants<br>N = 254,280 SNPs in polygenic score |
|---|---|---|---|---|
| Polygenic score | 1.27 (1.03, 1.57) | 1.28 (1.04, 1.59) | 1.27 (1.03, 1.57) | 1.19 (0.81, 1.77) |
| *C9orf72* repeat positive | 22.06 (2.73, 2859.55) | 20.82 (2.57, 2699.45) | - | 20.29 (2.52, 2627.47) |
| Family history of ALS | 29.14 (3.65, 3774.64) | 29.06 (3.63, 3767.08) | - | 28.06 (3.58, 3620.42) |
| Age (10-year increase) | 1.06 (0.87, 1.3) | 1.06 (0.87, 1.29) | 1.06 (0.87, 1.3) | 1.07 (0.88, 1.3) |
| Sex (male) | 1.57 (1.05, 2.36) | 1.58 (1.05, 2.38) | 1.57 (1.05, 2.36) | 1.52 (1.02, 2.29) |

SNP, single nucleotide polymorphism

*Supplementary Table S5. Genes mapping to the ALS polygenic score are enriched for Gene Ontology (GO) biologic processes.*

| GO | Pathway term | # of genes annotated to pathway | # of genes annotated to pathway significant in polygenic score | P-value for enrichment | Benjamini-Hochberg adjusted p-value for enrichment | Gene symbols for significant genes |
|---|---|---|---|---|---|---|
| GO:0030182 | neuron differentiation | 1357 | 28 | 8.74E-07 | 0.002196 | KIF5A,UNC13A,RAP1A,NFASC,WNT7A,REST,GRID2,FSTL4,TENM2,HDAC9,GLI3,ADCY1,CREB3L2,DSCAML1,RORA,PLXNB2,CHL1,TNIK,OSTN,PALLD,SEMA5A,CSMD3,KDM4C,PTPRD,DIP2B,MMD,APP,RUNX1 |
| GO:0120039 | plasma membrane bounded cell projection morphogenesis | 659 | 18 | 2.34E-06 | 0.002196 | KIF5A,UNC13A,NFASC,WNT7A,FSTL4,GLI3,ADCY1,CD44,DSCAML1,PLXNB2,CHL1,TNIK,OSTN,PALLD,SEMA5A,PTPRD,DIP2B,APP |
| GO:0048858 | cell projection morphogenesis | 663 | 18 | 2.55E-06 | 0.002196 | KIF5A,UNC13A,NFASC,WNT7A,FSTL4,GLI3,ADCY1,CD44,DSCAML1,PLXNB2,CHL1,TNIK,OSTN,PALLD,SEMA5A,PTPRD,DIP2B,APP |
| GO:0120036 | plasma membrane bounded cell projection organization | 1528 | 29 | 2.90E-06 | 0.002196 | KIF5A,UNC13A,RAP1A,NFASC,WNT7A,ABLIM2,GRID2,NEK1,FSTL4,TENM2,GLI3,ADCY1,CREB3L2,CD44,DSCAML1,DYNLL2,PARVB,PLXNB2,TANC1,CHL1,TNIK,OSTN,PALLD,SEMA5A,CSMD3,PTPRD,DIP2B,CDH13,APP |
| GO:0032990 | cell part morphogenesis | 679 | 18 | 3.56E-06 | 0.002196 | KIF5A,UNC13A,NFASC,WNT7A,FSTL4,GLI3,ADCY1,CD44,DSCAML1,PLXNB2,CHL1,TNIK,OSTN,PALLD,SEMA5A,PTPRD,DIP2B,APP |
| GO:0030030 | cell projection organization | 1568 | 29 | 4.81E-06 | 0.002196 | KIF5A,UNC13A,RAP1A,NFASC,WNT7A,ABLIM2,GRID2,NEK1,FSTL4,TENM2,GLI3,ADCY1,CREB3L2,CD44,DSCAML1,DYNLL2,PARVB,PLXNB2,TANC1,CHL1,TNIK,OSTN,PALLD,SEMA5A,CSMD3,PTPRD,DIP2B,CDH13,APP |
| GO:0032989 | cellular component morphogenesis | 766 | 19 | 4.89E-06 | 0.002196 | KIF5A,UNC13A,NFASC,WNT7A,MOBP,FSTL4,GLI3,ADCY1,CD44,DSCAML1,PLXNB2,CHL1,TNIK,OSTN,PALLD,SEMA5A,PTPRD,DIP2B,APP |
| GO:0000902 | cell morphogenesis | 1004 | 22 | 6.03E-06 | 0.002196 | KIF5A,UNC13A,NFASC,WNT7A,FSTL4,GLI3,ADCY1,DNMBP,CD44,DSCAML1,PARVB,PLXNB2,CHL1,TNIK,OSTN,PALLD,SEMA5A,SOX17,PTPRD,TEK,DIP2B,APP |
| GO:0048699 | generation of neurons | 1501 | 28 | 6.18E-06 | 0.002196 | KIF5A,UNC13A,RAP1A,NFASC,WNT7A,REST,GRID2,FSTL4,TENM2,HDAC9,GLI3,ADCY1,CREB3L2,DSCAML1,RORA,PLXNB2,CHL1,TNIK,OSTN,PALLD,SEMA5A,CSMD3,KDM4C,PTPRD,DIP2B,MMD,APP,RUNX1 |

| | | | | | | |
|---|---|---|---|---|---|---|
| GO:0048812 | neuron projection morphogenesis | 645 | 17 | 7.32E-06 | 0.002196 | KIF5A,UNC13A,NFASC,WNT7A,FSTL4,GLI3,ADCY1,DSCAML1,PLXNB2,CHL1,TNIK,OSTN,PALLD,SEMA5A,PTPRD,DIP2B,APP |
| GO:0022008 | neurogenesis | 1613 | 29 | 8.33E-06 | 0.002196 | KIF5A,UNC13A,RAP1A,NFASC,WNT7A,MOBP,REST,GRID2,FSTL4,TENM2,HDAC9,GLI3,ADCY1,CREB3L2,DSCAML1,RORA,PLXNB2,CHL1,TNIK,OSTN,PALLD,SEMA5A,CSMD3,KDM4C,PTPRD,DIP2B,MMD,APP,RUNX1 |
| GO:0000904 | cell morphogenesis involved in differentiation | 723 | 18 | 8.44E-06 | 0.002196 | KIF5A,NFASC,WNT7A,FSTL4,GLI3,ADCY1,DSCAML1,PARVB,PLXNB2,CHL1,TNIK,PALLD,SEMA5A,SOX17,PTPRD,TEK,DIP2B,APP |
| GO:0048666 | neuron development | 1109 | 23 | 8.89E-06 | 0.002196 | KIF5A,UNC13A,RAP1A,NFASC,WNT7A,GRID2,FSTL4,TENM2,GLI3,ADCY1,CREB3L2,DSCAML1,PLXNB2,CHL1,TNIK,OSTN,PALLD,SEMA5A,CSMD3,PTPRD,DIP2B,APP,RUNX1 |
| GO:0031175 | neuron projection development | 977 | 21 | 1.34E-05 | 0.003085 | KIF5A,UNC13A,RAP1A,NFASC,WNT7A,GRID2,FSTL4,GLI3,ADCY1,CREB3L2,DSCAML1,PLXNB2,CHL1,TNIK,OSTN,PALLD,SEMA5A,CSMD3,PTPRD,DIP2B,APP |
| GO:0007268 | chemical synaptic transmission | 683 | 17 | 1.54E-05 | 0.003085 | KIF5A,UNC13A,RAP1A,GRM7,WNT7A,GRID2,CLCN3,AKAP12,ADCY1,KCNQ3,GRIK4,ATXN3,DLGAP4,CHRM3,PTPRD,GIPC1,APP |
| GO:0098916 | anterograde trans-synaptic signaling | 683 | 17 | 1.54E-05 | 0.003085 | KIF5A,UNC13A,RAP1A,GRM7,WNT7A,GRID2,CLCN3,AKAP12,ADCY1,KCNQ3,GRIK4,ATXN3,DLGAP4,CHRM3,PTPRD,GIPC1,APP |
| GO:0007267 | cell-cell signaling | 1672 | 29 | 1.65E-05 | 0.003115 | KIF5A,UNC13A,RAP1A,GRM7,WNT7A,REST,GRID2,CLCN3,TMEM170B,AKAP12,GLI3,ADCY1,KCNQ3,FGF3,GRIK4,ATXN3,DLGAP4,PFKL,CHRM3,IL36G,TNIK,SEMA5A,MAP3K7,SOX17,PTPRD,TEK,GIPC1,APP,RUNX1 |
| GO:0099537 | trans-synaptic signaling | 690 | 17 | 1.75E-05 | 0.003125 | KIF5A,UNC13A,RAP1A,GRM7,WNT7A,GRID2,CLCN3,AKAP12,ADCY1,KCNQ3,GRIK4,ATXN3,DLGAP4,CHRM3,PTPRD,GIPC1,APP |
| GO:0099536 | synaptic signaling | 712 | 17 | 2.61E-05 | 0.004413 | KIF5A,UNC13A,RAP1A,GRM7,WNT7A,GRID2,CLCN3,AKAP12,ADCY1,KCNQ3,GRIK4,ATXN3,DLGAP4,CHRM3,PTPRD,GIPC1,APP |
| GO:0048667 | cell morphogenesis involved in neuron differentiation | 579 | 15 | 3.19E-05 | 0.005115 | KIF5A,NFASC,WNT7A,FSTL4,GLI3,ADCY1,DSCAML1,PLXNB2,CHL1,TNIK,PALLD,SEMA5A,PTPRD,DIP2B,APP |
| GO:0010646 | regulation of cell communication | 3440 | 46 | 3.37E-05 | 0.005156 | TNIP1,TBK1,UNC13A,BCAR3,RAP1A,GRM7,IRAK2,WNT7A,FOXP1,ZFYVE28,REST,GRID2,FSTL4,GCNT2,TMEM170B,AKAP12,GLI3,ADCY1,UBAP2,DUSP8,CD44,FGF3,GRIK4,RORA,RHOT2,ZDHHC7,GGNBP2,URI1,DLGAP4,SPATA2,PFKL,FAF1,TNIK,ADH7,SEMA5A,MAP3K7,SOX17,PTPRD,TEK,PIP5K1B,SRGAP1,NOD2,CDH13,GIPC1,APP,RUNX1 |

| | | | | | | |
|---|---|---|---|---|---|---|
| GO:1904861 | excitatory synapse assembly | 26 | 4 | 4.05E-05 | 0.005684 | WNT7A,GRID2,PLXNB2,PTPRD |
| GO:0023052 | signaling | 6731 | 74 | 4.28E-05 | 0.005684 | TNIP1,KIF5A,TBK1,UNC13A,BCAR3,RAP1A,NR1I3,NFASC,ZNF385B,GRM7,IRAK2,WNT7A,DOCK3,FOXP1,PLCXD2,ZFYVE28,SH3BP2,REST,GRID2,CLCN3,FSTL4,TENM2,GCNT2,TMEM170B,AKAP12,GLI3,ADCY1,CREB3L2,NSMAF,KCNQ3,UBAP2,FNBP1,GPR158,RAB18,DNMBP,DUSP8,CD44,FGF3,GRIK4,ITGBL1,ATXN3,RORA,RHOT2,ZDHHC7,GGNBP2,PITPNC1,URI1,DLGAP4,SPATA2,PFKL,PLXNB2,FAF1,CHRM3,IL36G,CHL1,FHIT,TNIK,OSTN,ADH7,SEMA5A,GMDS,MAP3K7,SOX17,PTPRD,TEK,PIP5K1B,OR10W1,SRGAP1,NOD2,CDH13,NXPH3,GIPC1,APP,RUNX1 |
| GO:0023051 | regulation of signaling | 3476 | 46 | 4.40E-05 | 0.005684 | TNIP1,TBK1,UNC13A,BCAR3,RAP1A,GRM7,IRAK2,WNT7A,FOXP1,ZFYVE28,REST,GRID2,FSTL4,GCNT2,TMEM170B,AKAP12,GLI3,ADCY1,UBAP2,DUSP8,CD44,FGF3,GRIK4,RORA,RHOT2,ZDHHC7,GGNBP2,URI1,DLGAP4,SPATA2,PFKL,FAF1,TNIK,ADH7,SEMA5A,MAP3K7,SOX17,PTPRD,TEK,PIP5K1B,SRGAP1,NOD2,CDH13,GIPC1,APP,RUNX1 |
| GO:0007409 | axonogenesis | 460 | 13 | 4.56E-05 | 0.005684 | KIF5A,NFASC,WNT7A,FSTL4,GLI3,ADCY1,DSCAML1,PLXNB2,CHL1,PALLD,SEMA5A,DIP2B,APP |
| GO:0007154 | cell communication | 6744 | 74 | 4.60E-05 | 0.005684 | TNIP1,KIF5A,TBK1,UNC13A,BCAR3,RAP1A,NR1I3,NFASC,ZNF385B,GRM7,IRAK2,WNT7A,DOCK3,FOXP1,PLCXD2,ZFYVE28,SH3BP2,REST,GRID2,CLCN3,FSTL4,TENM2,GCNT2,TMEM170B,AKAP12,GLI3,ADCY1,CREB3L2,NSMAF,KCNQ3,UBAP2,FNBP1,GPR158,RAB18,DNMBP,DUSP8,CD44,FGF3,GRIK4,ITGBL1,ATXN3,RORA,RHOT2,ZDHHC7,GGNBP2,PITPNC1,URI1,DLGAP4,SPATA2,PFKL,PLXNB2,FAF1,CHRM3,IL36G,CHL1,FHIT,TNIK,OSTN,ADH7,SEMA5A,GMDS,MAP3K7,SOX17,PTPRD,TEK,PIP5K1B,OR10W1,SRGAP1,NOD2,CDH13,NXPH3,GIPC1,APP,RUNX1 |
| GO:0050804 | modulation of chemical synaptic transmission | 405 | 12 | 5.77E-05 | 0.006771 | UNC13A,RAP1A,GRM7,WNT7A,GRID2,AKAP12,ADCY1,GRIK4,DLGAP4,PTPRD,GIPC1,APP |
| GO:0099177 | regulation of trans-synaptic signaling | 406 | 12 | 5.90E-05 | 0.006771 | UNC13A,RAP1A,GRM7,WNT7A,GRID2,AKAP12,ADCY1,GRIK4,DLGAP4,PTPRD,GIPC1,APP |
| GO:0034330 | cell junction organization | 699 | 16 | 7.54E-05 | 0.008346 | UNC13A,RAP1A,NFASC,WNT7A,REST,GRID2,PLEC,PLXNB2,TANC1,PTPRD,TEK,FMN1,CDH13,APP,RUNX1,ARVCF |
| GO:0007155 | cell adhesion | 1474 | 25 | 0.0001 | 0.010733 | TNIP1,NFASC,GRID2,TENM2,GCNT2,GLI3,CD44,DSCAML1,ITGBL1,ATXN3,PARVB,PLXNB2,FAF1,CHL1,PCDH10,PALLD,SEMA5A,PTPRD,TEK,FMN1,NOD2,CDH13,APP,RUNX1,ARVCF |

| | | | | | | |
|---|---|---|---|---|---|---|
| GO:0022610 | biological adhesion | 1481 | 25 | 0.000108 | 0.011196 | TNIP1,NFASC,GRID2,TENM2,GCNT2,GLI3,CD44,DSCAML1,ITGBL1,ATXN3,PARVB,PLXNB2,FAF1,CHL1,PCDH10,PALLD,SEMA5A,PTPRD,TEK,FMN1,NOD2,CDH13,APP,RUNX1,ARVCF |
| GO:1904889 | regulation of excitatory synapse assembly | 13 | 3 | 0.000113 | 0.011215 | WNT7A,GRID2,PTPRD |
| GO:1901888 | regulation of cell junction assembly | 195 | 8 | 0.000116 | 0.011215 | RAP1A,WNT7A,GRID2,PTPRD,TEK,FMN1,APP,RUNX1 |
| GO:0061564 | axon development | 507 | 13 | 0.000122 | 0.011215 | KIF5A,NFASC,WNT7A,FSTL4,GLI3,ADCY1,DSCAML1,PLXNB2,CHL1,PALLD,SEMA5A,DIP2B,APP |
| GO:0007399 | nervous system development | 2358 | 34 | 0.000122 | 0.011215 | KIF5A,UNC13A,RAP1A,NFASC,DEGS1,WNT7A,MOBP,REST,GRID2,FSTL4,TENM2,HDAC9,GLI3,ADCY1,CREB3L2,RAB18,DSCAML1,ATXN3,RORA,PLXNB2,CHRM3,CHL1,TNIK,OSTN,PALLD,SEMA5A,SOX17,CSMD3,KDM4C,PTPRD,DIP2B,MMD,APP,RUNX1 |
| GO:0009653 | anatomical structure morphogenesis | 2694 | 37 | 0.000155 | 0.01382 | KIF5A,UNC13A,BCAR3,RAP1A,NFASC,WNT7A,MOBP,FOXP1,GRID2,FSTL4,HDAC9,GLI3,ADCY1,DNMBP,CD44,FGF3,DSCAML1,RORA,ZDHHC7,PARVB,PLXNB2,TANC1,CHL1,TNIK,OSTN,PALLD,SEMA5A,SOX17,MMP16,PTPRD,TEK,DIP2B,FMN1,CDH13,ASXL3,APP,RUNX1 |
| GO:0031399 | regulation of protein modification process | 1628 | 26 | 0.000189 | 0.016384 | TNIP1,TBK1,BCAR3,RAP1A,IRAK2,DOCK3,ZFYVE28,ADCY1,DUSP8,CD44,FGF3,GGNBP2,URI1,PLXNB2,FAF1,TNIK,PHF1,MAP3K7,EIF2AK1,TEK,PPP2R2D,DIP2B,NOD2,MMD,UBA2,APP |
| GO:0051246 | regulation of protein metabolic process | 2631 | 36 | 0.000211 | 0.017831 | TNIP1,TBK1,BCAR3,RAP1A,IRAK2,WNT7A,DOCK3,RYBP,ZFYVE28,REST,SERPINB6,ADCY1,DUSP8,CD44,FGF3,ATXN2,ATXN3,GGNBP2,URI1,PLXNB2,FAF1,FHIT,TNIK,PHF1,MAP3K7,EIF2AK1,SOX17,TEK,PPP2R2D,DIP2B,NOD2,MMD,GIPC1,UBA2,APP,RUNX1 |
| GO:0098742 | cell-cell adhesion via plasma-membrane adhesion molecules | 273 | 9 | 0.000228 | 0.018343 | GRID2,TENM2,DSCAML1,PLXNB2,PCDH10,PALLD,PTPRD,CDH13,ARVCF |
| GO:0048468 | cell development | 2034 | 30 | 0.000228 | 0.018343 | KIF5A,UNC13A,RAP1A,NFASC,WNT7A,MOBP,FOXP1,REST,GRID2,FSTL4,TENM2,HDAC9,GLI3,ADCY1,CREB3L2,DSCAML1,PARVB,PLXNB2,CHL1,TNIK,OSTN,PALLD,SEMA5A,SOX17,CSMD3,PTPRD,TEK,DIP2B,APP,RUNX1 |
| GO:0001709 | cell fate determination | 41 | 4 | 0.000251 | 0.019445 | WNT7A,DSCAML1,NKX6-3,SOX17 |
| GO:0007411 | axon guidance | 277 | 9 | 0.000254 | 0.019445 | KIF5A,NFASC,GLI3,DSCAML1,PLXNB2,CHL1,PALLD,SEMA5A,APP |

| | | | | | | |
|---|---|---|---|---|---|---|
| GO:0097485 | neuron projection guidance | 278 | 9 | 0.000261 | 0.019505 | KIF5A,NFASC,GLI3,DSCAML1,PLXNB2,CHL1,PALLD,SEMA5A,APP |
| GO:0032268 | regulation of cellular protein metabolic process | 2473 | 34 | 0.000305 | 0.022247 | TNIP1,TBK1,BCAR3,RAP1A,IRAK2,WNT7A,DOCK3,RYBP,ZFYVE28,REST,SERPINB6,ADCY1,DUSP8,CD44,FGF3,ATXN2,ATXN3,GGNBP2,URI1,PLXNB2,FAF1,FHIT,TNIK,PHF1,MAP3K7,EIF2AK1,TEK,PPP2R2D,DIP2B,NOD2,MMD,GIPC1,UBA2,APP |
| GO:0007165 | signal transduction | 6239 | 67 | 0.000321 | 0.022914 | TNIP1,TBK1,UNC13A,BCAR3,RAP1A,NR1I3,ZNF385B,GRM7,IRAK2,WNT7A,DOCK3,FOXP1,PLCXD2,ZFYVE28,SH3BP2,GRID2,FSTL4,TENM2,GCNT2,TMEM170B,AKAP12,GLI3,ADCY1,CREB3L2,NSMAF,UBAP2,FNBP1,GPR158,RAB18,DNMBP,DUSP8,CD44,FGF3,GRIK4,ITGBL1,RORA,RHOT2,ZDHHC7,GGNBP2,PITPNC1,URI1,DLGAP4,SPATA2,PLXNB2,FAF1,CHRM3,IL36G,CHL1,FHIT,TNIK,OSTN,ADH7,SEMA5A,GMDS,MAP3K7,SOX17,PTPRD,TEK,PIP5K1B,OR10W1,SRGAP1,NOD2,CDH13,NXPH3,GIPC1,APP,RUNX1 |
| GO:0034329 | cell junction assembly | 425 | 11 | 0.000379 | 0.026303 | RAP1A,WNT7A,GRID2,PLEC,PLXNB2,PTPRD,TEK,FMN1,APP,RUNX1,ARVCF |
| GO:0099504 | synaptic vesicle cycle | 177 | 7 | 0.000391 | 0.026303 | UNC13A,RAP1A,WNT7A,CLCN3,ADCY1,DDC,GIPC1 |
| GO:0008038 | neuron recognition | 46 | 4 | 0.000393 | 0.026303 | DSCAML1,PALLD,SEMA5A,APP |
| GO:0010975 | regulation of neuron projection development | 428 | 11 | 0.000402 | 0.026357 | RAP1A,WNT7A,GRID2,FSTL4,CREB3L2,PLXNB2,TNIK,SEMA5A,CSMD3,PTPRD,DIP2B |
| GO:0021533 | cell differentiation in hindbrain | 20 | 3 | 0.000434 | 0.027851 | WNT7A,GRID2,RORA |
| GO:0051247 | positive regulation of protein metabolic process | 1529 | 24 | 0.000445 | 0.027997 | TNIP1,TBK1,BCAR3,RAP1A,IRAK2,WNT7A,DOCK3,REST,ADCY1,CD44,FGF3,ATXN3,PLXNB2,FAF1,TNIK,PHF1,MAP3K7,SOX17,TEK,DIP2B,NOD2,MMD,UBA2,APP |
| GO:0032270 | positive regulation of cellular protein metabolic process | 1447 | 23 | 0.000498 | 0.03078 | TNIP1,TBK1,BCAR3,RAP1A,IRAK2,WNT7A,DOCK3,REST,ADCY1,CD44,FGF3,ATXN3,PLXNB2,FAF1,TNIK,PHF1,MAP3K7,TEK,DIP2B,NOD2,MMD,UBA2,APP |
| GO:0009966 | regulation of signal transduction | 3074 | 39 | 0.000511 | 0.030946 | TNIP1,TBK1,BCAR3,RAP1A,IRAK2,WNT7A,FOXP1,ZFYVE28,FSTL4,GCNT2,TMEM170B,AKAP12,GLI3,UBAP2,DUSP8,CD44,FGF3,RORA,RHOT2,ZDHHC7,GGNBP2,URI1,DLGAP4,SPATA2,FAF1,TNIK,ADH7,SEMA5A,MAP3K7,SOX17,PTPRD,TEK,PIP5K1B,SRGAP1,NOD2,CDH13,GIPC1,APP,RUNX1 |
| GO:0035249 | synaptic transmission, glutamatergic | 90 | 5 | 0.000593 | 0.035258 | UNC13A,GRM7,GRID2,CLCN3,GRIK4 |

| | | | | | | |
|---|---|---|---|---|---|---|
| GO:0043410 | positive regulation of MAPK cascade | 525 | 12 | 0.000626 | 0.036569 | BCAR3,RAP1A,IRAK2,WNT7A,GCNT2,AKAP12,CD44,TNIK,MAP3K7,TEK,NOD2,APP |
| GO:0021697 | cerebellar cortex formation | 23 | 3 | 0.000662 | 0.037503 | WNT7A,GRID2,RORA |
| GO:0099003 | vesicle-mediated transport in synapse | 194 | 7 | 0.000675 | 0.037503 | UNC13A,RAP1A,WNT7A,CLCN3,ADCY1,DDC,GIPC1 |
| GO:0007215 | glutamate receptor signaling pathway | 53 | 4 | 0.000677 | 0.037503 | GRM7,GRID2,GRIK4,APP |
| GO:0120035 | regulation of plasma membrane bounded cell projection organization | 612 | 13 | 0.000737 | 0.040121 | RAP1A,WNT7A,GRID2,FSTL4,TENM2,CREB3L2,CD44,PLXNB2,TNIK,SEMA5A,CSMD3,PTPRD,DIP2B |
| GO:0001932 | regulation of protein phosphorylation | 1211 | 20 | 0.00075 | 0.040164 | TBK1,BCAR3,RAP1A,IRAK2,DOCK3,ZFYVE28,ADCY1,DUSP8,CD44,FGF3,GGNBP2,PLXNB2,FAF1,TNIK,MAP3K7,EIF2AK1,TEK,NOD2,MMD,APP |
| GO:2000300 | regulation of synaptic vesicle exocytosis | 55 | 4 | 0.00078 | 0.041052 | RAP1A,WNT7A,ADCY1,GIPC1 |
| GO:1901890 | positive regulation of cell junction assembly | 96 | 5 | 0.000795 | 0.0412 | WNT7A,GRID2,PTPRD,TEK,FMN1 |
| GO:0043408 | regulation of MAPK cascade | 708 | 14 | 0.000925 | 0.046805 | TNIP1,BCAR3,RAP1A,IRAK2,WNT7A,GCNT2,AKAP12,DUSP8,CD44,TNIK,MAP3K7,TEK,NOD2,APP |
| GO:0031401 | positive regulation of protein modification process | 1052 | 18 | 0.00094 | 0.046805 | TNIP1,TBK1,BCAR3,RAP1A,IRAK2,DOCK3,ADCY1,CD44,FGF3,TNIK,PHF1,MAP3K7,TEK,DIP2B,NOD2,MMD,UBA2,APP |
| GO:0031344 | regulation of cell projection organization | 629 | 13 | 0.000947 | 0.046805 | RAP1A,WNT7A,GRID2,FSTL4,TENM2,CREB3L2,CD44,PLXNB2,TNIK,SEMA5A,CSMD3,PTPRD,DIP2B |

*Supplementary Table S6. Genes mapping to the ALS polygenic score are enriched in Kyoto Encyclopedia of Genes and Genomes (KEGG) pathways.*

| KEGG annotation | Pathway term | # of genes annotated to pathway | # of genes annotated to pathway significant in polygenic score | P-value for enrichment | Benjamini-Hochberg adjusted p-value for enrichment | Gene symbols for significant genes |
|---|---|---|---|---|---|---|
| 4972 | Pancreatic secretion | 102 | 5 | 0.002309 | 0.471111 | RAP1A,PLA2G12A,ADCY1,CHRM3,PRSS3 |
| 71 | Fatty acid degradation | 43 | 3 | 0.006924 | 0.542835 | ACSL5,ACADS,ADH7 |
| 604 | Glycosphingolipid biosynthesis - ganglio series | 15 | 2 | 0.007983 | 0.542835 | B4GALNT1,ST6GALNAC3 |
| 4725 | Cholinergic synapse | 113 | 4 | 0.01947 | 0.647974 | ADCY1,CREB3L2,KCNQ3,CHRM3 |
| 4152 | AMPK signaling pathway | 120 | 4 | 0.023692 | 0.647974 | CREB3L2,PFKL,MAP3K7,PPP2R2D |
| 4728 | Dopaminergic synapse | 132 | 4 | 0.03214 | 0.647974 | KIF5A,DDC,CREB3L2,PPP2R2D |
| 51 | Fructose and mannose metabolism | 33 | 2 | 0.03613 | 0.647974 | PFKL,GMDS |
| 350 | Tyrosine metabolism | 36 | 2 | 0.042365 | 0.647974 | DDC,ADH7 |
| 4911 | Insulin secretion | 86 | 3 | 0.04353 | 0.647974 | ADCY1,CREB3L2,CHRM3 |

*Supplementary Table S7. Sample characteristics of the Spanish study used in replication and meta-analysis testing.*

| **Characteristic** | **Control** **N =** 2,756[1] | **ALS** **N =** 548[1] | **P-value**[2] |
|---|---|---|---|
| **ALS Polygenic score with C9 region removed** | -0.06 (-0.70, 0.63) | 0.01 (-0.59, 0.80) | 0.073 |
| **C9orf72 Repeat Status** | | | <0.001 |
| Negative | 2,756 (100%) | 538 (98%) | |
| Positive | 0 (0%) | 10 (1.8%) | |
| **Family History of ALS** | 0 (0%) | 43 (7.8%) | <0.001 |
| **Sex** | | | <0.001 |
| Male | 1,294 (47%) | 306 (56%) | |
| Female | 1,462 (53%) | 242 (44%) | |
| **Age** | 64 (51, 73) | 58 (48, 69) | <0.001 |
| **Genetic principal component 1** | -0.001 (-0.017, 0.016) | -0.007 (-0.016, 0.002) | <0.001 |
| **Genetic principal component 2** | 0.001 (-0.017, 0.017) | 0.002 (-0.012, 0.018) | 0.12 |
| **Genetic principal component 3** | 0.000 (-0.017, 0.017) | -0.003 (-0.018, 0.013) | 0.024 |
| **Genetic principal component 4** | 0.000 (-0.016, 0.017) | 0.000 (-0.016, 0.017) | 0.6 |
| **Genetic principal component 5** | -0.001 (-0.017, 0.016) | -0.001 (-0.018, 0.015) | >0.9 |

[1]Median (IQR); n (%)

[2]Wilcoxon rank sum test; Fisher's exact test; Pearson's Chi-squared test

**Figures**

*Supplementary Figure S1. Participant filtering based on A). genetic data quality control and B). inclusion criteria and data availability.*

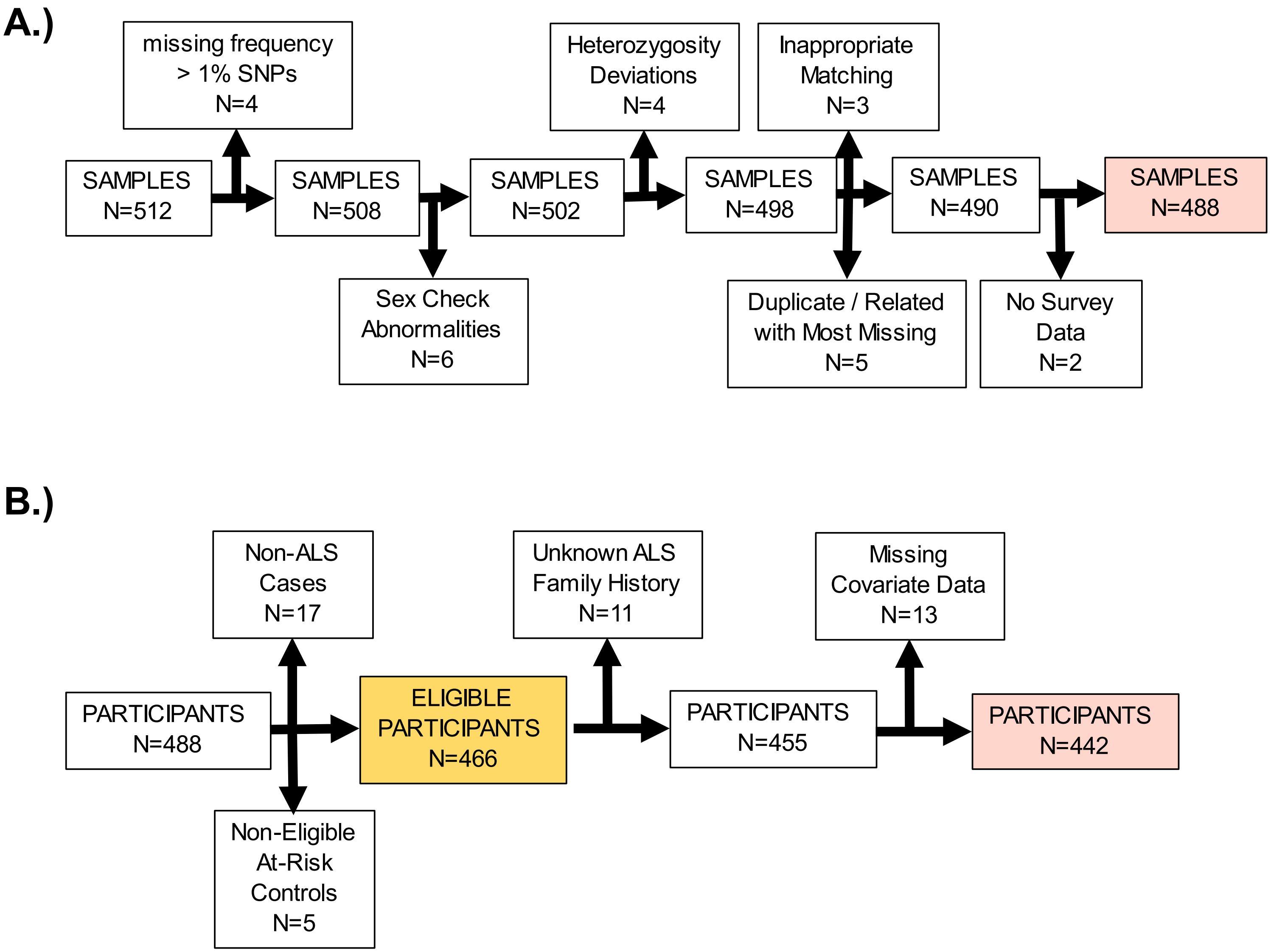

*Supplementary Figure S2. Single nucleotide polymorphism (SNP) filtering based on genetic data quality control assessments.*

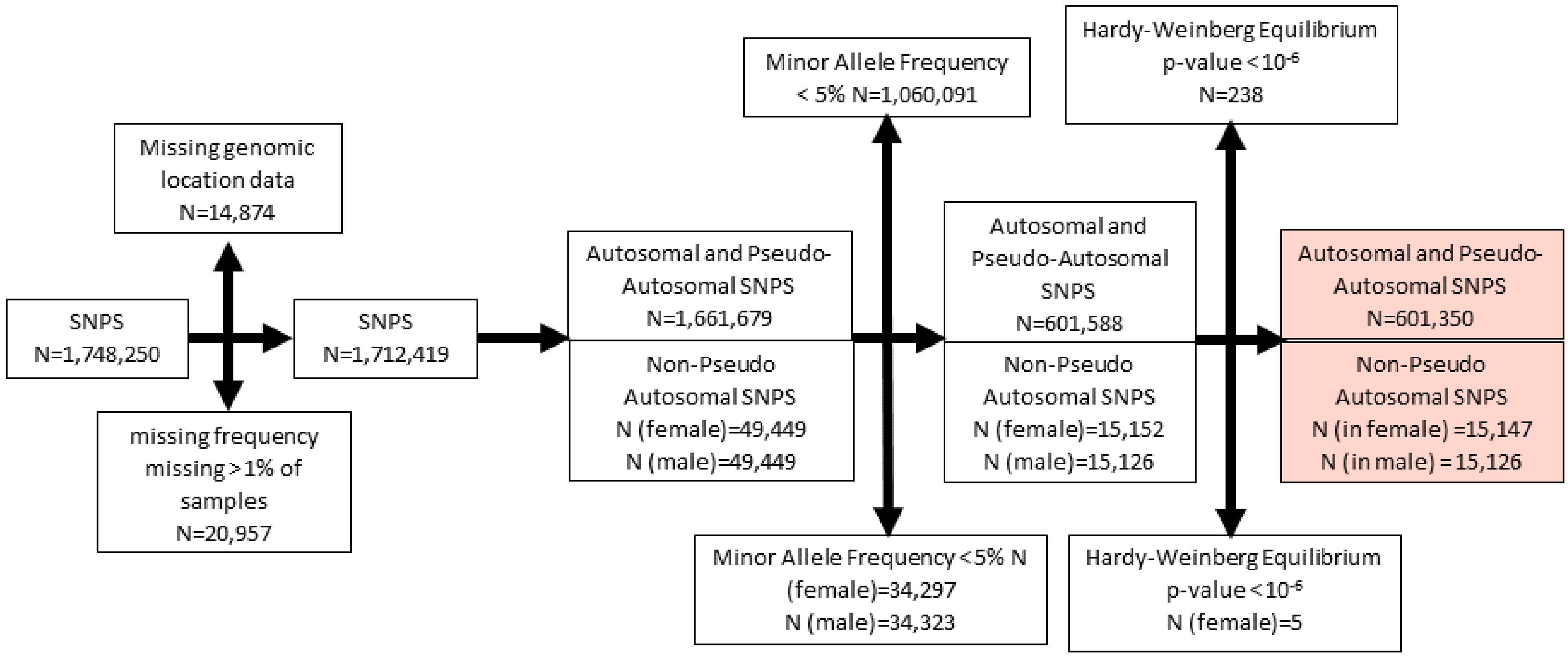

*Supplementary Figure S3. Post imputation single nucleotide polymorphism (SNP) filtering.*

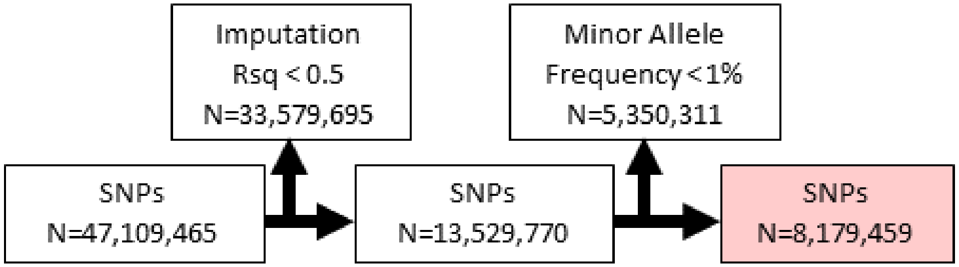

*Supplementary Figure S4. P-value threshold for single nucleotide polymorphism (SNP) inclusion in the ALS polygenic score without the C9orf72 region.*

P-value threshold represents the level of significance for SNPs observed in an ALS genome-wide association study (Nicholas et al. 2018).[9] Polygenic scores for ALS were calculated in an independent case-control sample (n = 208 controls, n=228 cases). The model fit reflects the percentage variance in ALS status explained by the polygenic score and the $-\log_{10}$(p-value) of this polygenic score with ALS status in this sample. The numbers above the bars represent the number of SNPs in polygenic score at given p-value threshold. The number inside the bars and color represent the p-value for polygenic score model fit. Polygenic score analysis shows the model fit is highest (p-value ~ $10^{-4}$ and has the lowest p-value threshold (p-value = 0.037) using 275 SNPs.

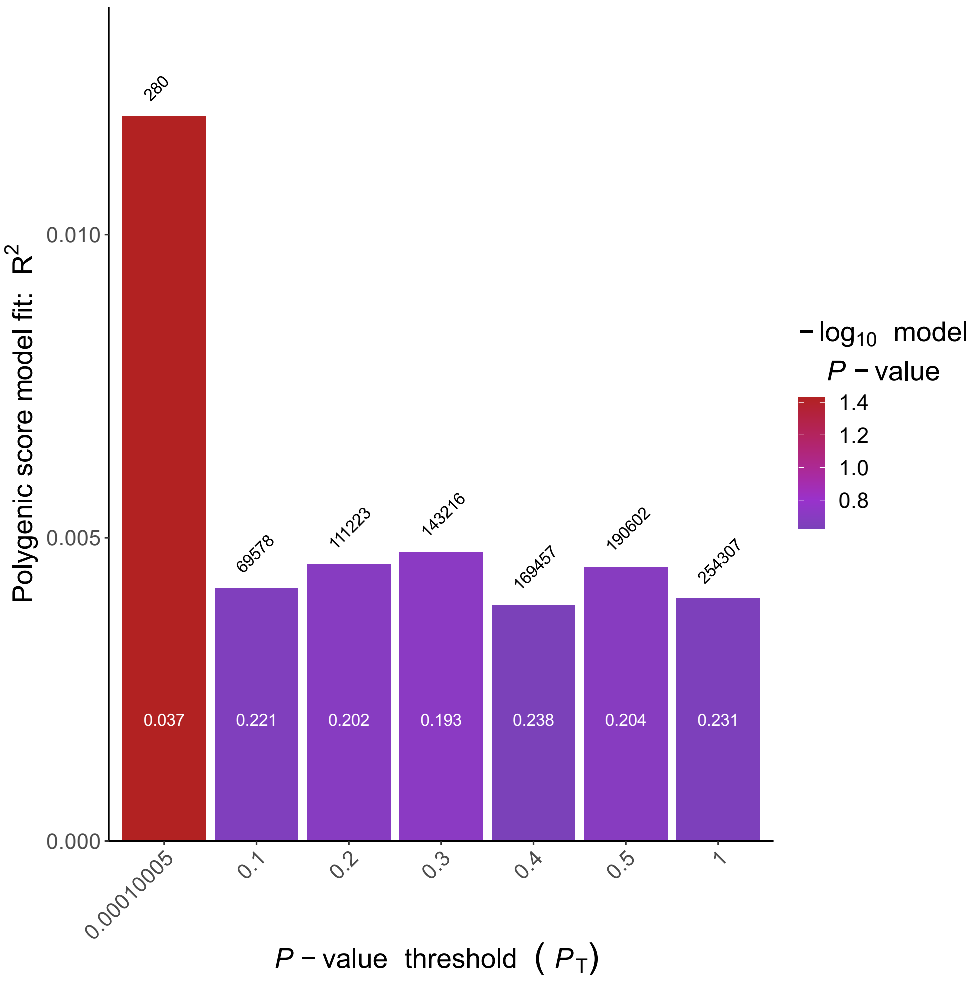

*Supplementary Figure S5. P-value threshold for single nucleotide polymorphism (SNP) inclusion in the ALS polygenic score containing the C9orf72 region.*
P-value threshold represents the level of significance for SNPs observed in an ALS genome-wide association study (Nicholas et al. 2018).[9] Polygenic scores for ALS are calculated in an independent case-control sample (n=208 controls, n = 228 cases). The model fit reflects the percentage variance in ALS status explained by the polygenic score and the -$\log_{10}$(p-value) of this polygenic score with ALS status in this sample. The numbers above the bars represent the number of SNPs in polygenic score at the given p-value threshold. The number inside the bars and color represent the p-value for polygenic score model fit. Polygenic score analysis shows the model fit is highest (p-value ~ $10^{-4}$ and has the lowest p-value threshold (p-value = 0.037) using 280 SNPs.

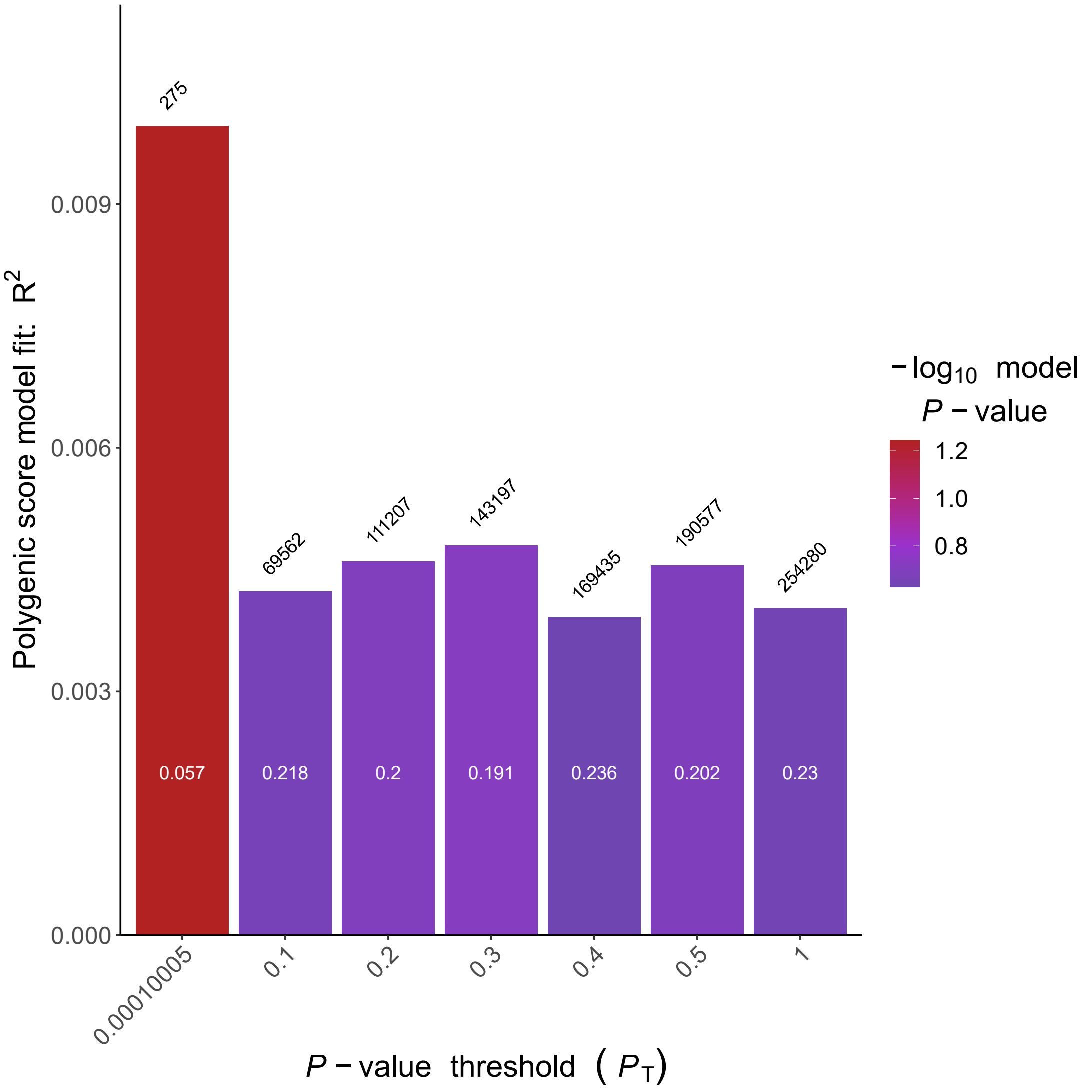

*Supplementary Figure S6. Distribution of ALS polygenic score by ALS case status.*
The polygenic score has been z-score standardized to be mean centered at zero with a standard deviation of one in the sample. The y-axis corresponds to the proportion of observations at that score. Blue represents the ALS case sample and red represents the control sample.

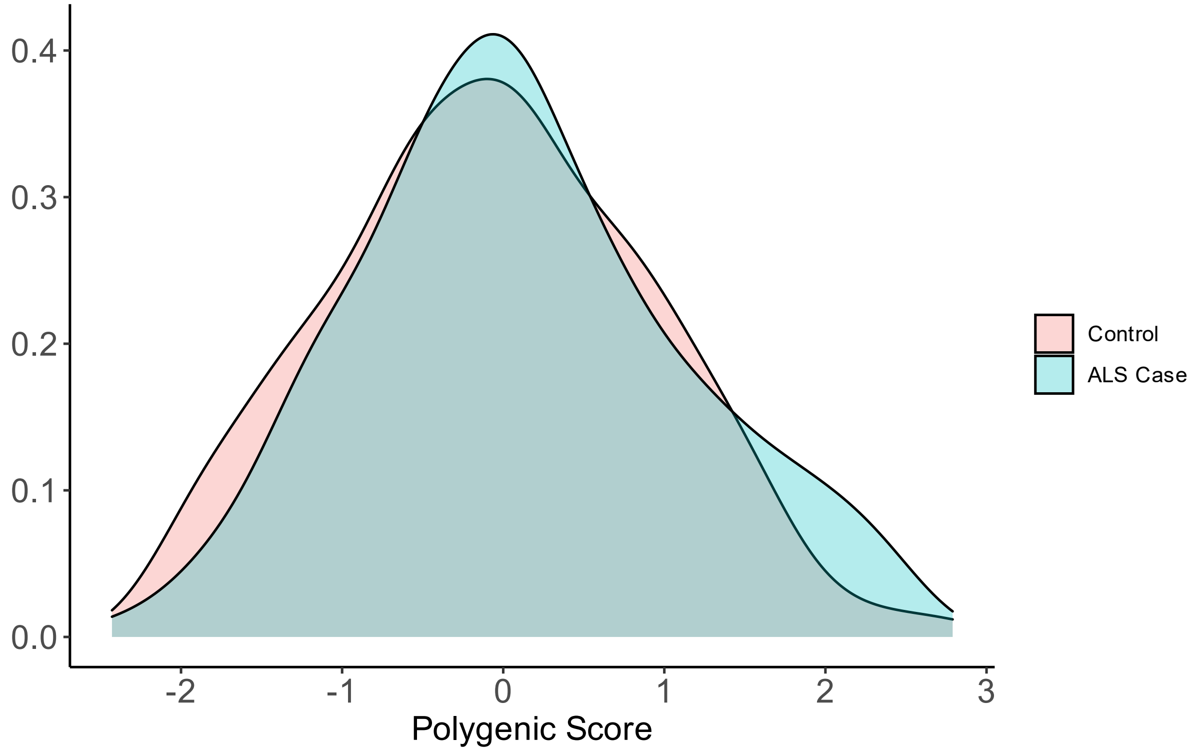

*Supplementary Figure S7. Receiver operator characteristic (ROC) curve with 5-fold cross validation predicted probabilities (n = 435).*

ROC for classifying ALS and control participants. The base model includes sex, age, and five genetic principal components and has an area under the curve (AUC) of 0.539 (blue). Adding family history to the base model increases the AUC to 0.588 (fam his; yellow). Adding *C9orf72* repeat expansion status in addition to family history increases the AUC to 0.603 (C9; red). Adding polygenic score (PGS; purple) in addition to family history and *C9orf72* repeat expansion status improves the AUC to 0.620.

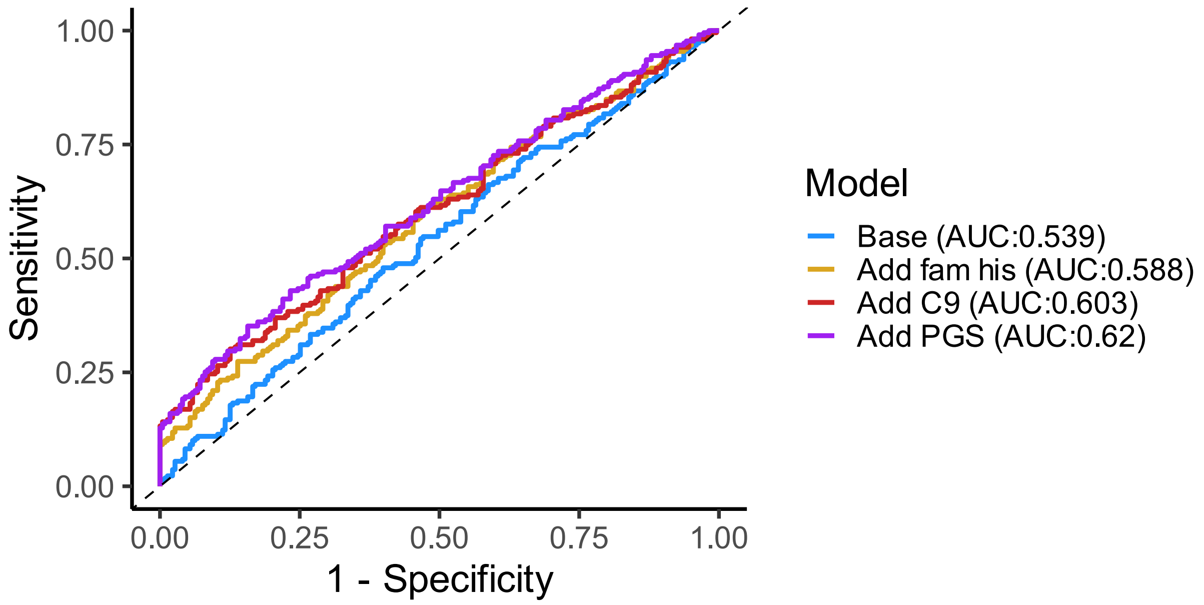

*Supplementary Figure S8. Receiver operator characteristic (ROC) curve, European ancestry only (n = 409).*
ROC curve for classifying ALS and control participants for the European ancestry sample only. The base model includes sex, age, and five genetic principal components and has an area under the curve (AUC) of 0.569 (blue). Adding family history to the base model increases the AUC to 0.611 (likelihood ratio test p-value = 0.42) (fam his; yellow). Further adding *C9orf72* repeat expansion status increases the AUC to 0.630 (likelihood ratio test p-value = 0.01) (C9; red). Next, adding polygenic score (PGS; purple) with the *C9orf72* region removed improves the AUC to 0.647 (likelihood ratio test p-value < 0.001).

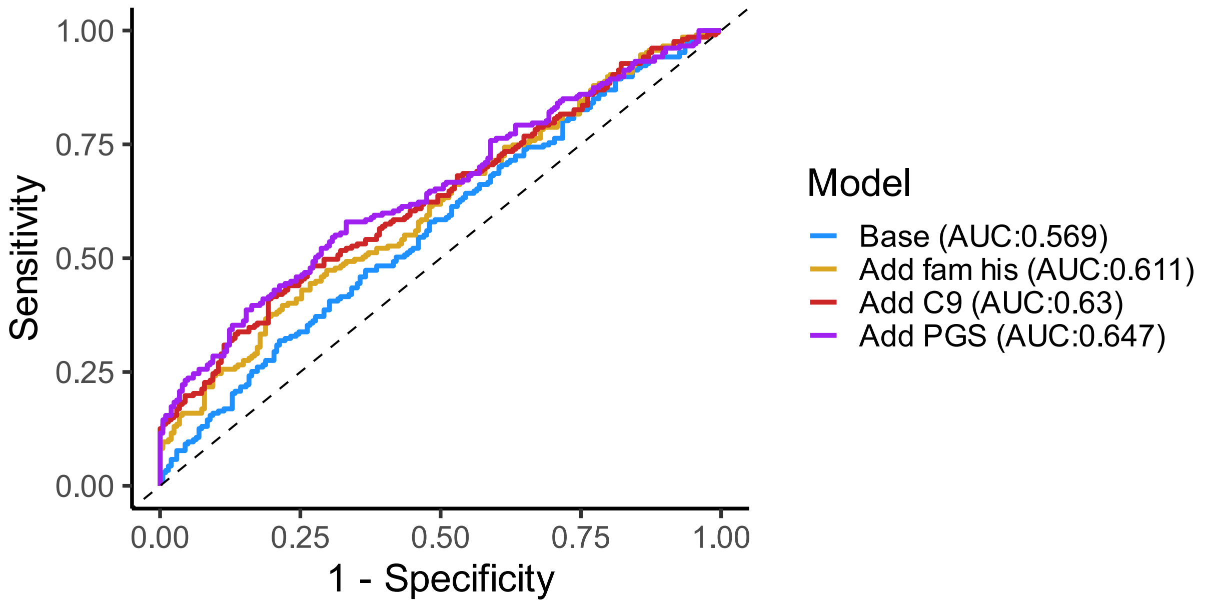

*Supplementary Figure S9. Meta-analysis and cohort level association of ALS polygenic score and ALS case status.*
Forest plot shows regression estimate odds ratio for one standard deviation increase in ALS polygenic score with 95% confidence interval. Meta-analysis in red, Spanish cohort in blue, Michigan cohort in green.

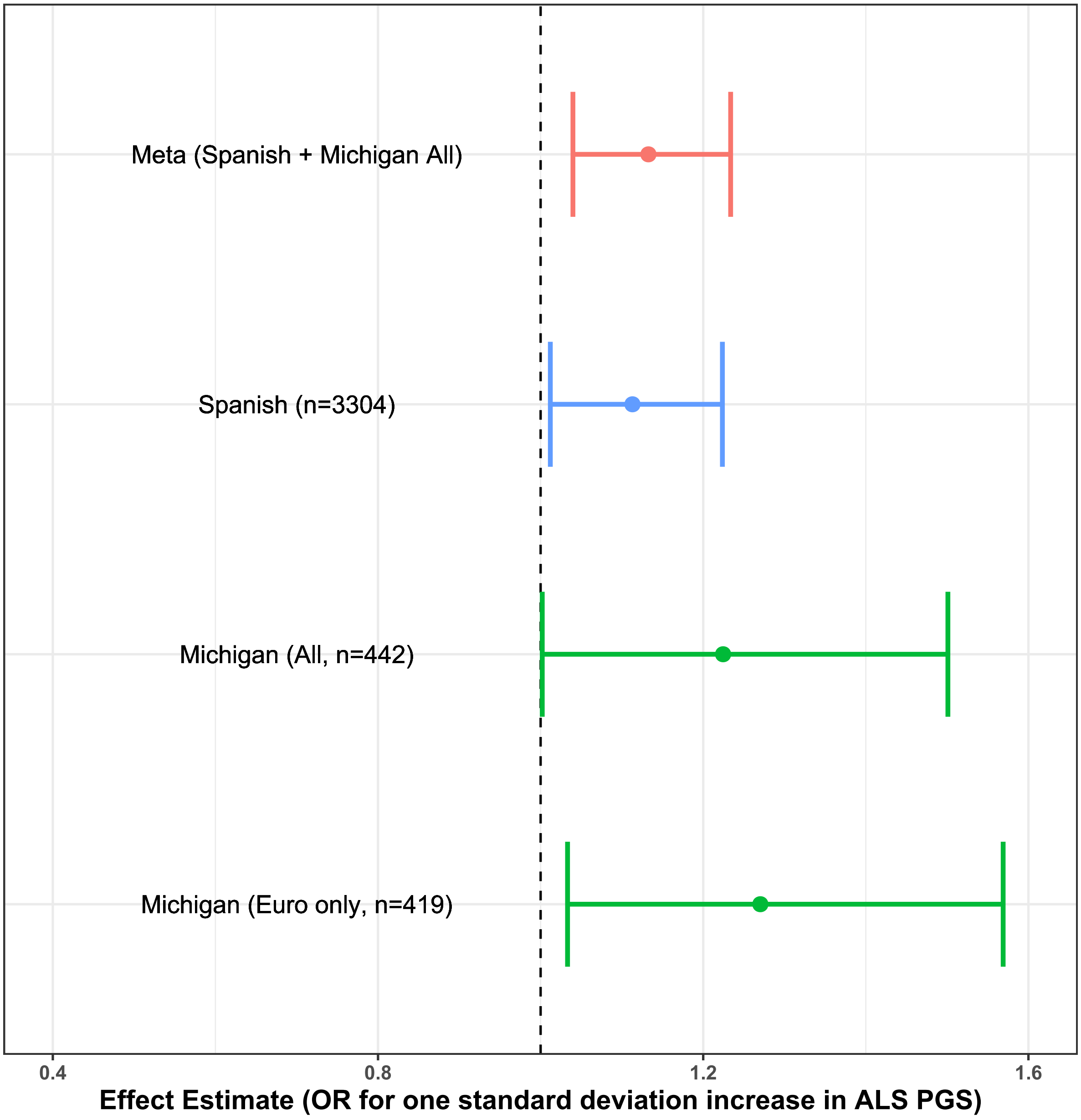
